# CSF proteomic analysis of semorinemab Ph2 trials in prodromal-to-mild (Tauriel) and mild-to-moderate (Lauriet) Alzheimer’s disease identifies distinct trial cell-type specific proteomic signatures

**DOI:** 10.1101/2024.04.11.24305670

**Authors:** Alyaa M. Abdel-Haleem, Ellen Casavant, Balazs Toth, Edmond Teng, Cecilia Monteiro, Nikhil J. Pandya, Casper C. Hoogenraad, Brad A. Friedman, Felix L. Yeh, Veronica G. Anania, Gloriia Novikova

## Abstract

Targeting of tau pathology has long been proposed as a potential therapeutic strategy for Alzheimer’s disease (AD). Semorinemab is a humanized IgG4 monoclonal antibody that binds to all known isoforms of full-length tau with high affinity and specificity. Semorinemab’s safety and efficacy have been studied in two Phase 2 randomized, double-blind, placebo-controlled, parallel-group clinical trials: Tauriel (prodromal-to-mild AD; NCT03289143) and Lauriet (mild-to-moderate AD; NCT03828747). CSF was collected from a subset of patients at baseline and after 49 or 73 weeks in Tauriel and baseline and after 49 or 61 weeks in Lauriet. We generated a large proteomics dataset, using more than 250 cerebrospinal fluid (CSF) samples and detecting more than 3500 proteins, to investigate the effects of semorinemab in each trial. Treatment-induced proteomic signatures were defined for each study as a set of proteins significantly elevated in the treatment arm in the respective study. Integration of the corresponding gene signatures with two independent brain single-nucleus RNA-seq datasets from AD and healthy aged controls revealed that Lauriet signature genes were enriched in microglial cells, while Tauriel signature genes were more broadly expressed across major brain cell types. Furthermore, the Lauriet trial gene signature was significantly upregulated in microglia from AD patients as compared to non-demented controls. The elevation of proteins such as CHI3L1 and GPNMB with treatment suggested an activated glial state. Taken together, this study utilizes a large CSF clinical proteomics dataset to assess the pharmacodynamic response of semorinemab and contributes to our understanding of how an anti-tau antibody influences disease-relevant pathophysiology in AD.

## Introduction

Alzheimer’s disease (AD) is a significant global health challenge marked by the relentless progression of cognitive decline and functional impairment. The neuropathological hallmarks of AD involve the accumulation of extracellular β-amyloid (Aβ) plaques and intracellular neurofibrillary tangles (NFTs) composed of tau protein (*1*). Tau pathology, in particular, correlates closely with cognitive deterioration, making it an attractive target for therapeutic intervention(*2*).

Semorinemab (also known as RO7105705, MTAU9937A, or RG6100) is a humanized IgG4 monoclonal antibody that targets the N-terminal domain of tau (amino acid residues 6-23) and binds to all known isoforms of full-length tau (*3*, *4*). Semorinemab was tested in two Phase 2 randomized, placebo-controlled clinical trials: the Tauriel study enrolled participants with prodromal-to-mild AD (50 to 80 years old, met criteria for mild cognitive impairment or dementia due to AD, and had a screening Mini-Mental Status Examination (MMSE) score between 20 and 30, inclusive, and Clinical Dementia Rating global score (CDR-GS) of 0.5 or 1) and the Lauriet study enrolled participants with mild-to-moderate AD (50 to 85 years old, met criteria for AD dementia, and had a screening MMSE score between 16 and 21, inclusive, and a CDR-GS of 1 or 2) (*3*, *5*). In the Tauriel study, semorinemab (in doses up to 8,100 mg IV Q4W) did not show differences between treatment arms and placebo in the progression of multiple clinical endpoints, including in the Clinical Dementia Rating-Sum of Boxes (CDR-SB) and the 13-item version of the Alzheimer’s Disease Assessment Scale-Cognitive Subscale (ADAS-Cog13). In the Lauriet study, patients treated with semorinemab (4,500 mg IV Q4W) demonstrated a significantly slower cognitive decline relative to those treated with placebo as measured by the ADAS-Cog11 (one of the two co-primary efficacy endpoints); however, semorinemab had no effect on the other co-primary outcome measure, the Alzheimer’s Disease Cooperative Study-Activities of Daily Living (ADCS-ADL), or on either of the secondary outcome measures (MMSE and CDR-SB). Due to the differences in clinical outcomes observed in Lauriet and Tauriel, we decided to explore the pharmacodynamic response to semorinemab in these two studies.

Large-scale proteomics approaches have recently been utilized to investigate the underlying biology of AD, with many of them focusing on brain tissues (*6*, *7*).

Cerebrospinal fluid (CSF) is a clear liquid surrounding the brain and spinal cord, and it has been broadly utilized as an accessible biofluid for biomarker discovery in AD (*8*, *9*). To this end, amidst the differences in clinical findings between the Tauriel and Lauriet studies, we utilized deep CSF proteomic profiling approaches to unravel the molecular intricacies underlying semorinemab’s differential impact. Utilizing an unbiased data analysis approach, trial signatures were defined for each study as the set of proteins that were significantly elevated with semorinemab treatment. We integrated the corresponding gene signatures with single-nucleus RNA-seq (snRNA-seq) datasets from AD and control brains to investigate their likely cellular origins and relevance to the disease. The results suggest that the Lauriet trial signature genes are enriched in microglial cells, while the Tauriel trial signature genes are expressed broadly across brain-resident cells. Interestingly, genes encoding proteins from the Lauriet trial signature are significantly upregulated in AD brains as compared to aged controls and correlated with a continuous pseudo-progression score, which is a continuous measure of pathology burden, suggesting that these proteins are positively correlated with pathology burden and a likely activated glial state. Taken together, this study provides valuable insights into molecular responses to semorinemab, potentially contributing to our understanding of semorinemab and other anti-tau antibody therapies.

## Results

### Generation of the CSF proteomic dataset

To characterize the baseline CSF proteome of AD patients as well as proteomic responses to semorinemab, CSF was collected from a subset of patients at baseline and after 49 and/or 73 weeks in Tauriel and baseline and after 49 or 61 weeks for Lauriet (Supplementary Table 1). A discovery data independent acquisition mass spectrometry (DIA-MS) approach was used to identify and quantify proteins in the CSF (Methods). More than 3500 proteins were measured, with an average of 3028 proteins per sample, demonstrating a broad dynamic range with protein intensities spanning 7 orders of magnitude (Figure 1, Supplementary Figure 1A-B, Methods).

**Figure 1.**
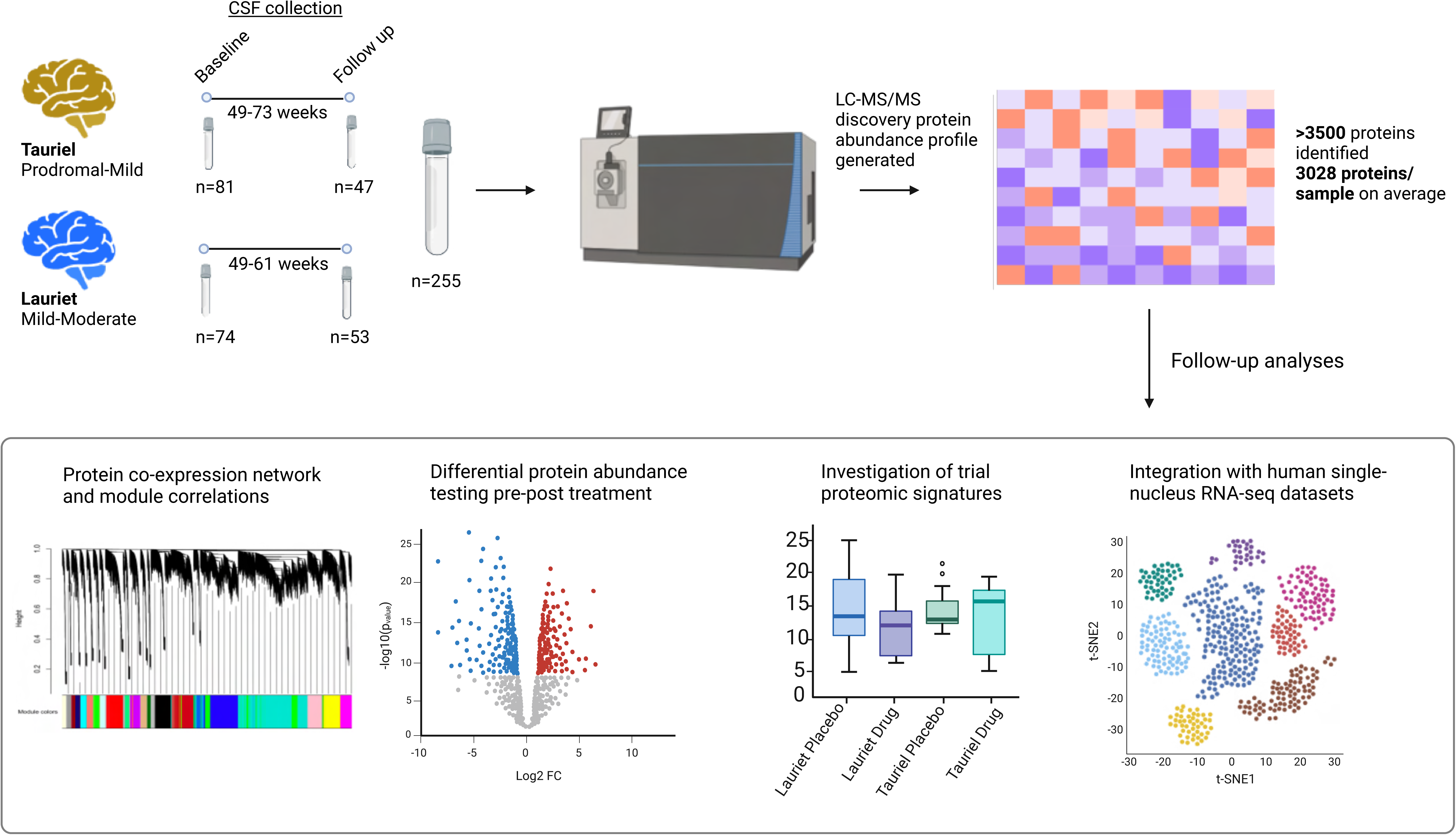
Study workflow. CSF samples from Lauriet and Tauriet participants were collected. The Tauriel study contained 81 samples at baseline and 47 samples at follow-up, while the Lauriet study contained 74 samples at baseline and 53 at follow-up, resulting in a total of 255 samples (Supplementary Table 1). LC-MS/MS CSF protein abundances were generated, containing more than 3500 proteins with an average of 3028 proteins per sample. Follow-up analysis included protein network construction using WGCNA, differential abundance testing, investigation of the trial proteomic signatures, and integration with human RNA-seq datasets.

### Network analysis reveals protein modules correlated with baseline clinical characteristics and cell-type markers

To investigate the structure of our CSF proteomic dataset, a network analysis using a weighted gene coexpression network analysis (WGCNA) framework was performed. Broadly, WGCNA is able to detect groups or clusters of highly correlated proteins, which by nature of their co-expression are more likely to reflect shared biological pathways (*10*). For each such cluster (module) identified by WGCNA, an eigenprotein is calculated, which summarizes the protein expression profile of the module and can be further correlated to traits of interest (*10*). To investigate the AD CSF proteome, only baseline samples were used (n=155) to avoid confounding effects with the treatment group to perform WGCNA analysis, which resulted in 15 modules ranging in size from the largest (*n* = 1429 proteins) to the smallest (*n* = 12 proteins) (Figure 2A, Methods). Module correlations with baseline clinical characteristics were then assessed, including CDR-SB, ADAS-Cog (ADAS-Cog11 and ADAS-Cog13 in Lauriet and Tauriel studies, respectively), ADCS-ADL, and MMSE, as well as imaging outcomes, including tau PET imaging using Genentech Tau Probe 1 (GTP1)(*11*) across a whole cortical gray region of interest (GTPWCG) and whole brain volumetric MRI (MRIWB). Several modules (modules 4, 8, 9, 12, 13, and 15) significantly correlated with at least one of the traits tested (Spearman cor ≥ 0.1, nominal P-value ≤ 0.05) (Figure 2B). For example, a positive correlation was observed between module 12 and GTPWCG (Figure 2B). MAPT (encodes the tau protein) resided in this module and its protein levels significantly correlated with the tau PET signal (Figure 2D). Interestingly, YWHAZ and YWHAG (encode 14-3-3Z and 14-3-3G proteins, respectively), proteins that were previously shown to be significantly upregulated in the CSF of AD patients, belonged to the same module and also showed a significant correlation with the tau PET signal (data for YWHAZ shown in Figure 2D)(*8*). We identified multiple modules correlating to individual measures of cognitive ability or global function; for example, a significant negative correlation was observed between module 13 and MMSE (Figure 2B). Interestingly, brain levels of two proteins residing in that module, GAPDH (cor. with MMSE = -0.46, P = 1.5e-9) and CNP (cor. with MMSE = -0.31, P = 1.6e-4), were previously shown to be associated with cognitive trajectory defined using longitudinal MMSE data using two independent cohorts(*12*). However, we did not identify a module correlating with multiple complementary measures of cognitive impairment (e.g., ADAS-Cog11/13, MMSE), likely due to multiple factors, including the high variability inherent to these clinical assessments, the moderate to mild correlations observed between these measures, and the limited variability in these measures at baseline within the AD patient cohort.

**Figure 2.**
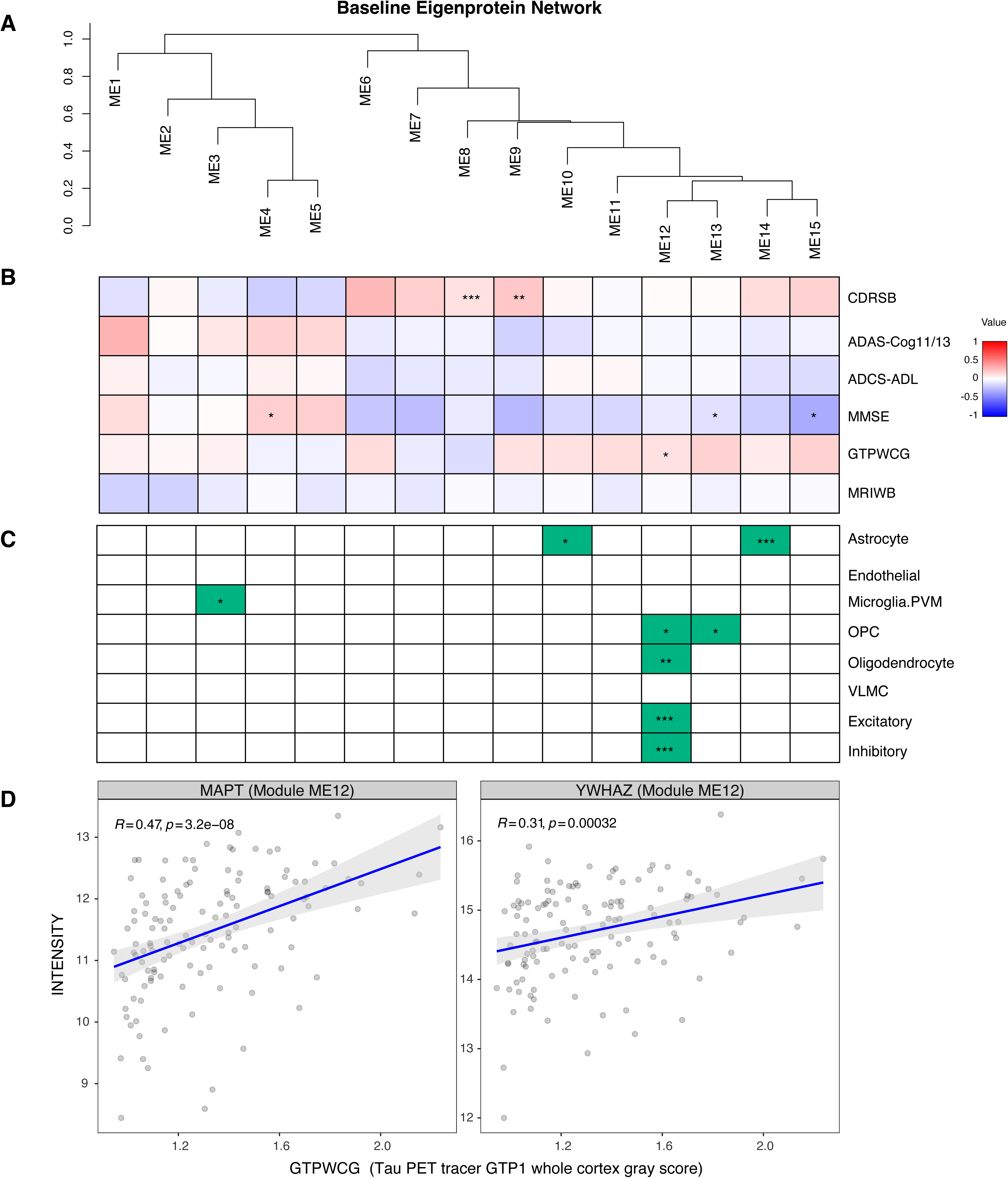
WGCNA network analysis of the baseline CSF proteome. **A.** Dendrogram showing WGCNA results of the baseline CSF proteome. **B.** Spearman correlation analysis of the module eigenproteins with baseline clinical characteristics: Clinical Dementia Rating-Sum of Boxes (CDRSB), Alzheimer’s Disease Assessment Scale-Cognitive Subscale (ADAS-Cog11/13), Alzheimer’s Disease Cooperative Study-Activities of Daily Living (ADCS-ADL), Mini-Mental State Examination (MMSE), tau PET imaging using Genentech Tau Probe 1 (GTP1) across a whole cortical gray region of interest (GTPWCG) and whole brain volumetric MRI (MRIWB). The strengths of positive (red) and negative (blue) correlations are shown in the heatmap with asterisks highlighting significant correlations (unadjusted P-value <= 0.05) with an absolute correlation value of at least 0.1. **C.** The significance of cell-type enrichment for each module was calculated using a hypergeometric test to assess the significance of the overlap between module proteins and cell-type specific markers from the SEA-AD dataset. Green boxes and asterisks denote all significant correlations (unadjusted P-value <= 0.05). OPC, Oligodendrocyte precursor cells; Microglia.PVM, Microglia and Perivascular macrophages; VLMC, vascular leptomeningeal cells. Excitatory and Inhibitory labels refer to Vip (vasoactive intestinal peptide-expressing interneurons) and L2/3 neuronal layer as the most abundant excitatory and inhibitory neuronal subtypes in the SEA-AD dataset. For panels A and B, if a P-value <= 0.05, it is flagged by one star (*), a P-value <= 0.01 is flagged by two stars (**), and a P-value <= 0.001 is flagged with three stars (***). **D.** Correlation plots between log2 of protein intensity for MAPT and YWHAZ on the y-axis and tau PET signal (GTPWCG) on the x-axis.

Given that the CSF proteome is at least in part reflective of the brain proteome, enrichment of module proteomes in brain cell-type specific markers derived from Seattle Alzheimer’s disease Brain Cell Atlas (SEA-AD) post-mortem brain snRNA-seq dataset from AD patients and healthy aged donors was assessed(*13*) (Methods). Interestingly, 5 out of 15 modules contain a significant number of cell-type specific markers, including a broad enrichment of module 12, which contains the largest number of proteins whose corresponding genes were highly expressed in inhibitory and excitatory neurons as well as oligodendrocyte precursor cells (OPC) and oligodendrocytes, including MBP, TAC3, LAMA3, COL5A2, ADAMTS3 and TNR (Figure 2C). Some modules exhibited more specific enrichment: for example, module 3 was significantly enriched in microglial markers, including HLA-DRA, while module 10 was significantly enriched in astrocytic markers, including AQP4 and GFAP (Figure 2C). Overall, this unbiased network-based approach led to the identification of groups of correlated CSF proteins, suggesting their putative cell types of origin and providing the modules’ potential links to baseline clinical outcomes.

### Proteins detected in the CSF likely originate from brain cells as well as other tissues

CSF surrounds the CNS, is derived from plasma, and is secreted by the choroid plexuses, hence its proteome likely reflects both peripheral and CNS dynamics(*14*). Given that the CSF originates from plasma, we reasoned that a considerable portion of the proteome likely originates from the periphery. Indeed, when the expression of genes corresponding to the proteins detected in the CSF proteomics dataset was mapped to the Genotype-Tissue Expression (GTEx) project RNA-seq data, enrichment of brain tissues as well as other non-CNS tissues, including liver, spleen, and whole blood was observed, supporting this hypothesis (Figure 3A). Seeing a predictable enrichment of brain tissues in our CSF proteome using GTEx RNA-seq data, we wanted to investigate the overlap between detected proteins in our baseline CSF dataset and in a recently published AD brain proteomic dataset (*8*). We found that approximately 61% of proteins detected in the CSF were also detected in the brain, suggesting that these proteins are likely, at least in part, derived from the brain (data not shown).

**Figure 3.**
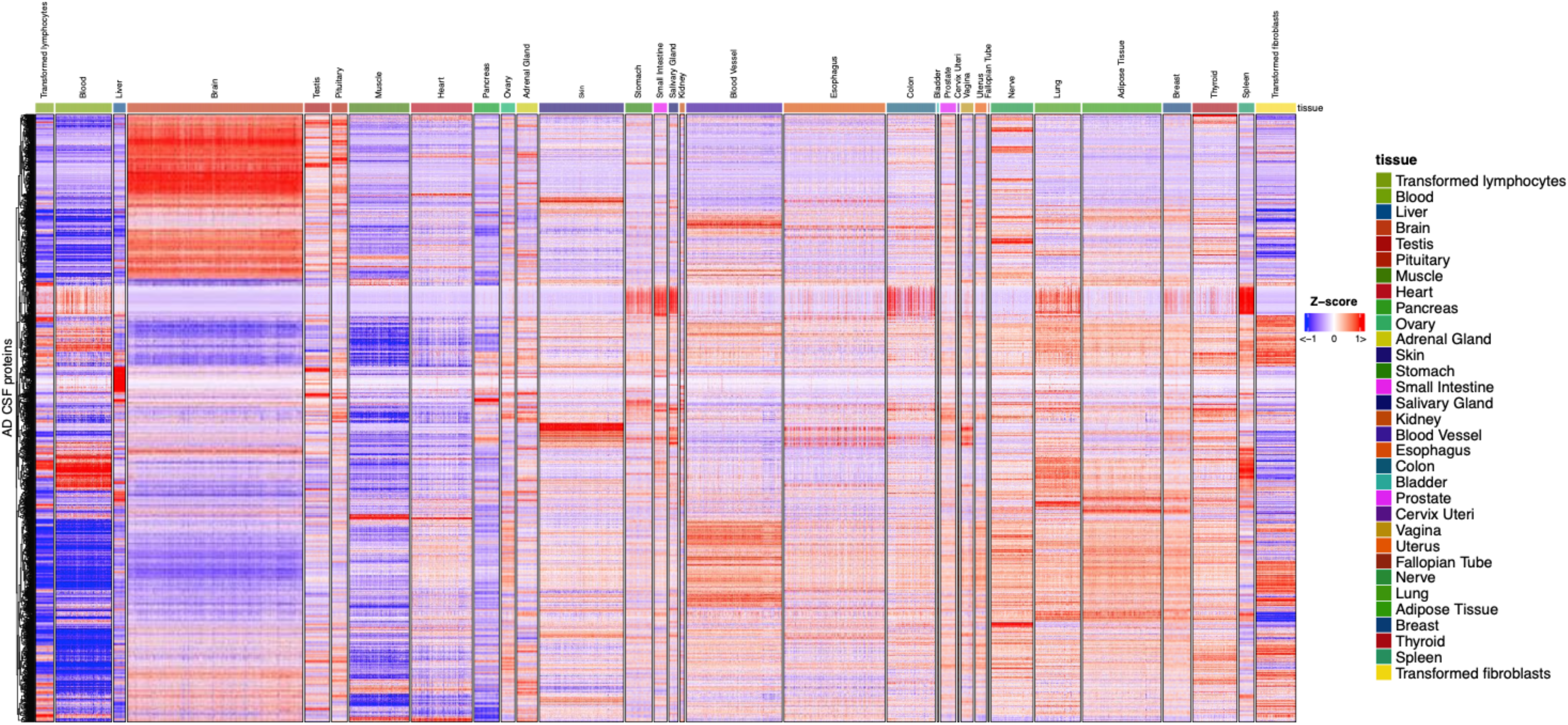
AD CSF proteome is enriched in brain tissues as well as other non-CNS tissues. Heatmap showing row-normalized *Z* scores for genes encoding proteins detected in the CSF proteome mapped across tissues from the GTEx project bulk RNA-seq dataset. Each row represents the normalized gene expression level of the gene encoding the protein detected in the AD CSF proteome. Each column represents a donor.

We next sought to examine the likely cell types of origin of our detected proteins, focusing specifically on brain-resident cell types. We discovered that along with detecting genes that were broadly expressed in multiple cell types, a range of cell-type enriched genes, such as ADAMTS3 (excitatory neurons), SHISA8 (inhibitory neurons), MBP (oligodendrocytes), GFAP (astrocytes), CSF1R (microglia) and others were detectable, suggesting that their corresponding proteins detected in the CSF could be produced by these cells (Figures 4 and Supplementary Figure 2A). Furthermore, these findings were confirmed in an independent brain snRNA-seq dataset from AD and control brain tissues, reproducibly demonstrating the detection of gene expression of corresponding CSF proteins in human brain cells (Supplementary Figure 2B)(*15*). Since cells residing in CSF can also secrete proteins, the CSF proteomics dataset was integrated with a snRNA-seq from human CSF(*16*). Indeed, a subset of genes corresponding to detected CSF proteins were expressed by CSF-resident cells, including AXL and CXCL12 in CSF myeloid cells (Supplementary Figure 2C). Taken together, these results demonstrate that a significant portion of the CSF proteins is detected in the brain proteome and expressed by various brain-resident cell types, suggesting that they likely, at least in part, originate from the brain.

**Figure 4.**
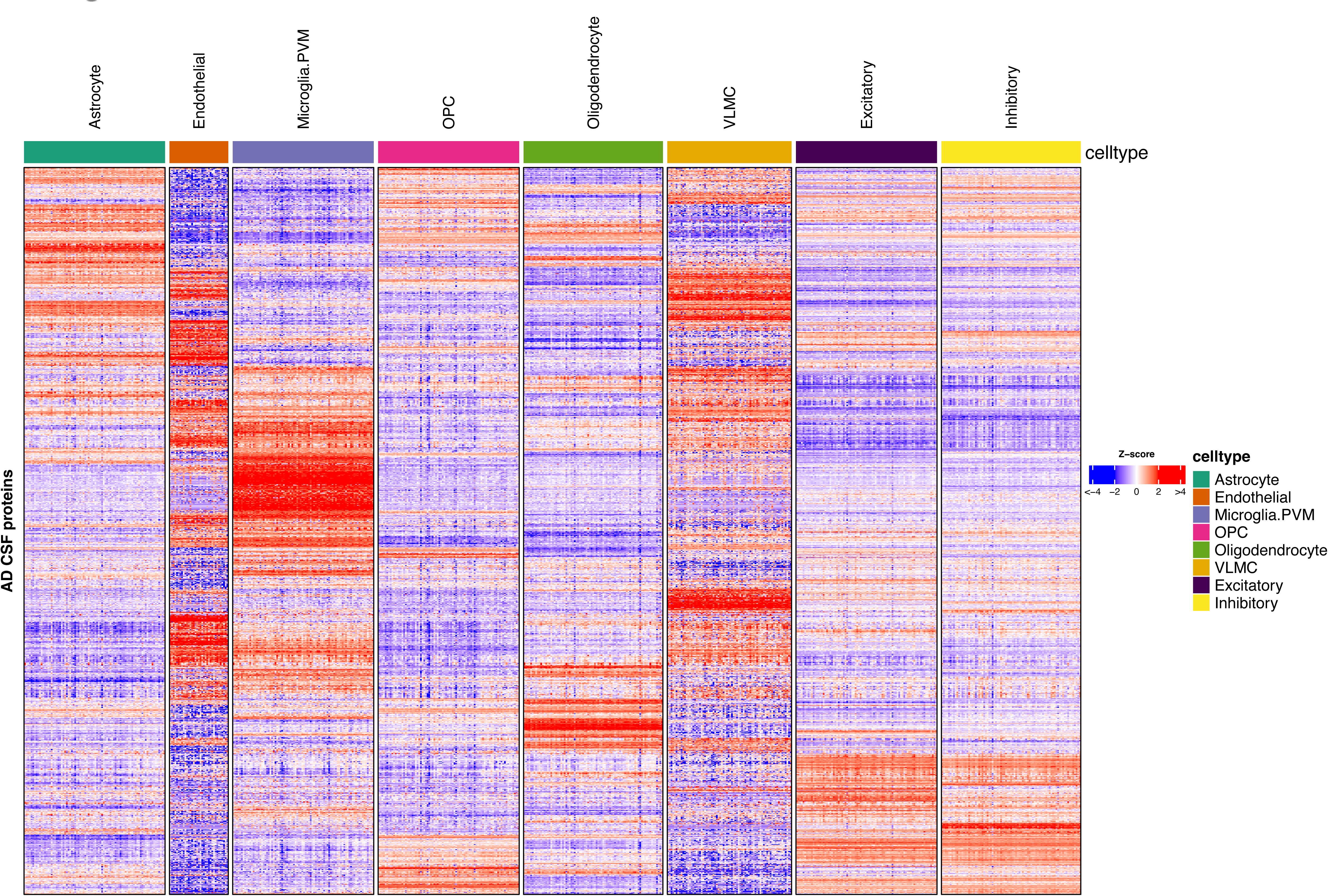
Expression of genes encoding AD CSF proteins across brain-resident cell types. Heatmap showing row-normalized Z scores for genes encoding AD CSF proteins across pseudobulk profiles from the SEA-AD brain snRAN-seq dataset. Each row represents the normalized gene expression of the gene encoding a protein detected in the CSF proteome. Each column represents a pseudobulk profile by donor and cell type from the SEA-AD dataset (Methods). OPC, Oligodendrocyte precursor cells; Microglia.PVM, Microglia and Perivascular macrophages; VLMC, vascular leptomeningeal cells. Excitatory and Inhibitory labels refer to Vip (vasoactive intestinal peptide-expressing interneurons) and L2/3 neuronal layer as the most abundant excitatory and inhibitory neuronal subtypes in the SEA-AD dataset. Other excitatory and inhibitory subtypes showed similar expression patterns (Supplementary Figure 2A).

### Defining Lauriet and Tauriel trial proteomic signatures

To understand the response of the CSF proteome to semorinemab in both Lauriet and Tauriel trials, we performed differential abundance analyses of the proteome pre- and post-treatment (Methods). In order to eliminate potential dose effects, only samples from the 4,500 mg arm in the Tauriel study were evaluated. We were able to identify 27 and 37 proteins significantly altered in Lauriet and Tauriel semorinemab treatment groups, respectively (baseline to follow-up comparison, FDR < 10%, after filtering for missingness and removing immunoglobulin G proteins (IgG)) (Figure 5A, Supplementary Figure 3A-B). Similar to prior reports, a downregulation of MAPT in both trials was observed (Figure 5A)(*3*, *5*). Despite this, a limited consistency in proteins significantly altered in both trials was found with CHI3L1, CHI3L2, and CXCL10 being significantly upregulated in Lauriet and trending in Tauriel (Figure 5A). To investigate potential treatment-induced cell-type specific activation profiles, we focused on proteins that are upregulated in follow-up vs. baseline groups in each study, resulting in 14 and 33 signature proteins associated with Lauriet and Tauriel trials, respectively (Figure 5B). As expected from our signature construction strategy, the Lauriet protein signature was significantly upregulated within the Lauriet semorinemab treatment group only, and not in placebo groups or in any of the Tauriel groups (Supplementary Figure 3C). Although there were no individual proteins significantly changed in placebo groups in both trials, we did observe that the overall Tauriel protein signature was significantly upregulated in both the placebo and semorinemab treatment groups (Supplementary Figure 3C).

**Figure 5.**
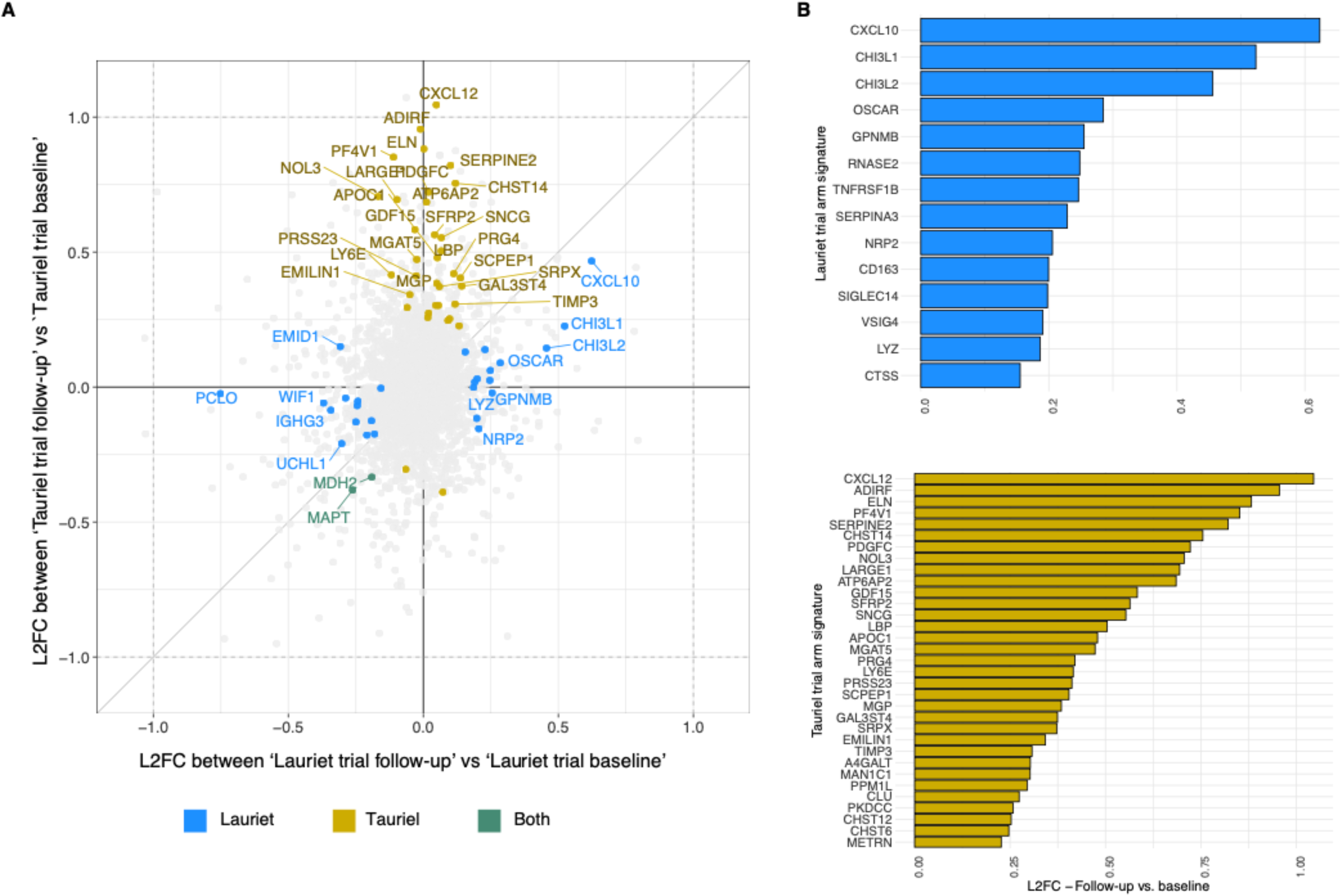
**A.** 4-way comparison of log fold changes in Lauriet (x-axis) and Tauriel (y-axis) trial arm protein signatures. Additional filtration criteria and removal of IgGs have also been applied to define the trial arm drug signatures (see text and Methods for details). Each point represents a protein colored by whether its corresponding FDR was < 0.1 in one or both trials (golden for Tauriel, blue for Lauriet, and cyan for both). **B.** Bar plot of Log2 fold changes (L2FCs) comparing follow-up and baseline treatment arms for individual proteins within the Lauriet and Tauriel trial signatures. LFCs are based on differential protein abundance analysis using limma (see text/methods for details).

However, the effect size in the semorinemab treatment group (0.38) was more than twice as large as the effect size observed in the placebo group (0.17) (Supplementary Figure 3C).

### Lauriet trial gene signature is enriched in microglial cells, while the Tauriel trial gene signature is expressed more broadly

To identify the likely cell type driving the trial proteomic signatures, we estimated the average expression of Lauriet and Tauriel signatures in brain cell types in the SEA-AD dataset(*13*). We identified genes that encode proteins that constitute the Lauriet and Tauriel proteomic signatures, which we refer to as Lauriet and Tauriel trial signature genes. Strikingly, a strong significant enrichment of genes preferentially expressed by microglial cells in Lauriet trial signature genes was observed (Figure 6A, Supplementary Figures 4, 5A, and 6). Conversely, the Tauriel signature genes were broadly expressed across major brain cell types (Figure 6A, Supplementary Figures 5A, and 6). These observations were validated in an independent brain snRNA-seq dataset (Morabito et al) from AD and control tissues, showing a strong microglia gene expression of the Lauriet trial signature (Supplementary Figure 7A)(*15*). Taken together, these findings suggest that CSF proteins that were significantly upregulated with treatment in the Lauriet trial potentially originate from microglial cells in the brain, while proteins that were upregulated with treatment in the Tauriel trial were likely derived from multiple cell types.

**Figure 6.**
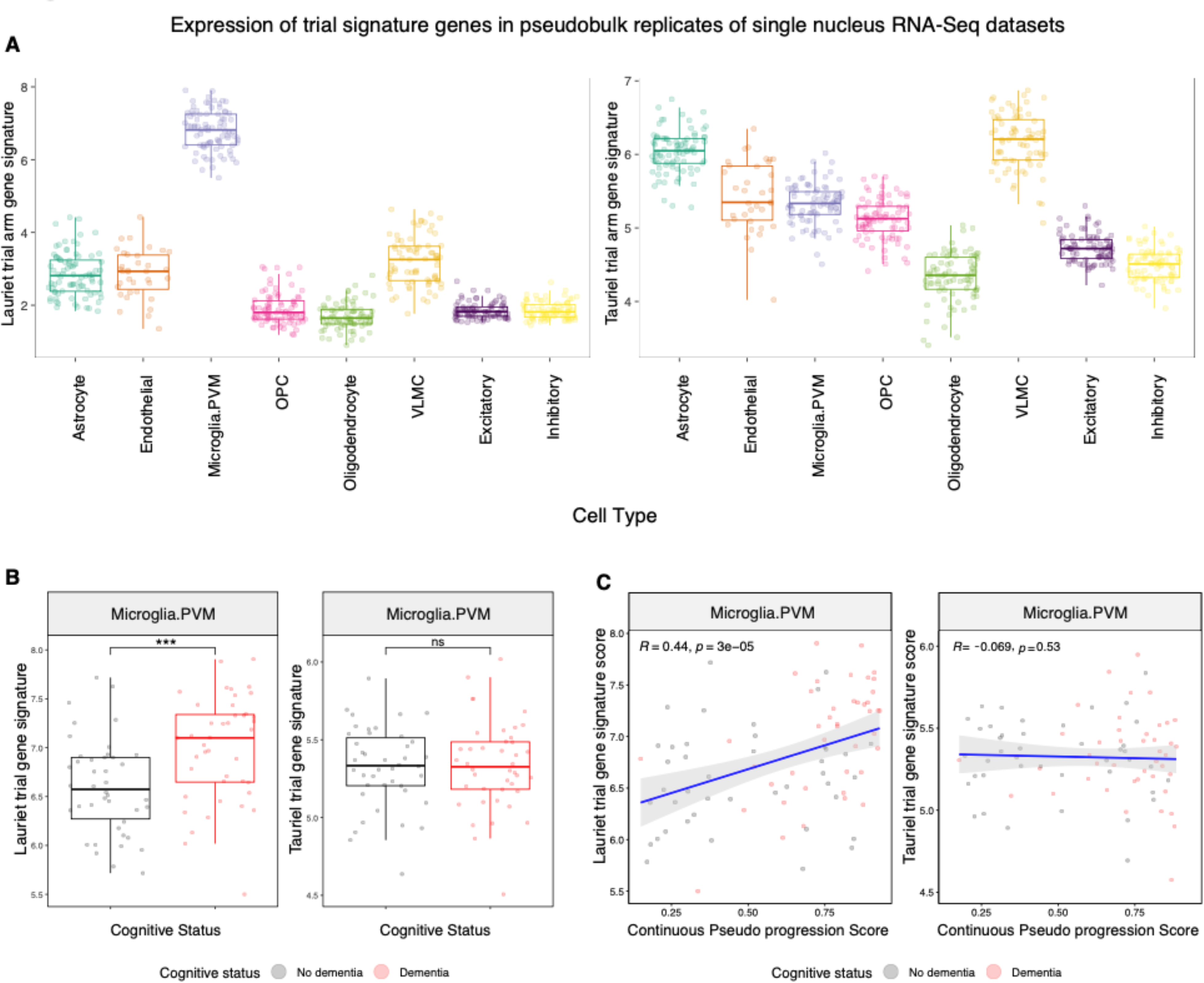
Lauriet trial gene signature is enriched in microglia and is correlated with disease pseudoprogression. **A.** Distribution of Lauriet and Tauriel gene set scores computed across pseudo bulk profiles of cell types from the SEA-AD brain dataset (see Methods). OPC, Oligodendrocyte precursor cells; Microglia.PVM, Microglia and Perivascular macrophages; VLMC, vascular leptomeningeal cells. Excitatory and Inhibitory labels refer to Vip (vasoactive intestinal peptide-expressing interneurons) and L2/3 neuronal layer as the most abundant excitatory and inhibitory neuronal subtypes in the SEA-AD dataset. Other excitatory and inhibitory subtypes showed similar gene set scores (Supplementary Figure 5) **B.** Distribution of Lauriet and Tauriel gene set signatures in demented and non-demented subjects from the SEA-AD dataset. P-values were derived using the Wilcoxon test. **C.** Correlation plots between the Lauriet and Tauriel gene signature score and a continuous pseudoprogression score (CPS), obtained for each donor from the SEA-AD brain dataset.

### A subset of Lauriet signature proteins is elevated in AD brains

Several CSF proteins that were upregulated in the semorinemab arm in the Lauriet trial, including GPNMB and SERPINA3, were previously reported as being upregulated at the mRNA level in mouse models of neurodegeneration as well as in human AD brains(*17*, *18*). To test if Lauriet signature genes are upregulated in AD brains, we utilized the SEA-AD snRNA-seq dataset to quantify the average expression of Lauriet signature genes between these two groups in microglial cells specifically(*13*). Indeed, significantly higher expression of this gene set in AD brains was observed, suggesting that some of the proteins elevated in the Lauriet trial are already elevated in diseased brains (Figure 6B, Supplementary Figure 5B). Conversely, Tauriel trial-associated genes were similarly expressed in microglia from AD and control brains (Figure 6B). These findings have been replicated in an independent dataset (Morabito et al), exhibiting a reproducible induction of Lauriet trial-associated genes in microglia in AD brains relative to controls (Supplementary Figure 7B)(*15*). Next, we asked whether proteins significantly upregulated in the Lauriet trial potentially correlate with disease progression. To assess this, we utilized a pseudo-progression score that is calculated using image-based quantitative neuropathology and provided within the SEA-AD dataset for each donor (*13*). Lauriet signature genes were, on average, positively correlated with the pseudo-progression score, suggesting that they change dynamically with the pathological burden of the brain (Figure 6C). These findings suggest that a subset of Lauriet signature genes are elevated in AD brains, potentially reflecting a microglial activation state that responds to accumulating pathology.

## Discussion

The observation that, unlike amyloid plaques, the burden of neurofibrillary tangles correlates with cognitive decline in AD has sparked increasing interest in anti-tau therapies, with several tau antibodies entering clinical trials in recent years(*2*, *19*). Although treatment with semorinemab did not slow the clinical progression of AD patients enrolled in the Tauriel study, the Lauriet study met one of the co-primary endpoints (ADAS-Cog11), demonstrating a 42.2 % reduction in decline (vs placebo)(*5*).

In this study, we aimed to leverage omics approaches to provide further molecular insights into our clinical findings, deepening our understanding of anti-tau approaches for the treatment of AD.

To this end, a mass-spectrometry-based CSF proteomics dataset on a subset of Tauriel and Lauriet participants was generated, which, to our knowledge, is the first CSF proteomics study that provides insight into the pharmacodynamic response of an anti-tau antibody. More than 3500 proteins were measurable in the CSF, which is a comparable or greater number of proteins than detected in recently published AD CSF proteome datasets (*8*, *9*, *20*). By integrating the CSF proteomics dataset with tissue-specific bulk RNA-seq data from GTEx as well as single-nucleus RNA-seq data from post-mortem brains, we were able to show that although, predictably, a portion of the CSF proteome is likely derived from the periphery, a substantial proportion of detected proteins likely originate from the brain based on their gene expression in brain-resident cell types.

Through this multi-omic analysis, we discovered that Tauriel and Lauriet signatures are largely distinct, sharing only 2 proteins (amongst them an expected reduction in MAPT)(*3*, *5*). This observation could be driven by the difference in patient populations enrolled in the studies: Tauriel study included participants with prodromal to mild AD, while the Lauriet population had more advanced cognitive impairment, diagnosed with mild to moderate AD. Hence, the distinct proteomic responses to treatment with semorinemab could be driven by an interaction of drug treatment with pathology burden. Indeed, a recent meta-analysis of AD proteomic studies highlighted numerous proteomic changes evident only in late-stage AD as compared to early AD(*21*).

We also report an intriguing finding that trial signatures demonstrate differences in cell-type specificity, with the Lauriet trial signature genes strongly enriched in microglial cells in the brain, while Tauriel signature genes show a broader expression pattern.

Furthermore, we observed that Lauriet signature genes were upregulated in AD brains as compared to aged controls in two independent post-mortem snRNA-seq datasets, and their expression was correlated with neuropathological burden in the brain. These findings suggest that the Lauriet trial signature could be reflective of the activated microglial state that is induced by more advanced disease pathology (as compared to the Tauriel trial) and augmented by semorinemab.

Microglia are innate myeloid immune cells and are resident macrophages of the brain(*22*). One of their main functions as phagocytes is the clearance of brain debris both during the development of the CNS and under pathological conditions(*22*). Microglia have taken center stage in AD research in recent years after being strongly implicated in AD pathogenesis through the discovery of rare and non-coding variants affecting genes highly and/or exclusively expressed in microglia in the brain(*23–25*). In the context of AD pathology, microglia are hypothesized to perform many functions, some of which include phagocytosis of cellular debris and protein aggregates, the release of cytokines, and the creation of a protective barrier around plaques, preventing further neurotoxic damage(*25*, *26*). This process is dependent on TREM2 in both mouse models of AD and in humans carrying an R47H mutation in the gene, impairing TREM2 ligand binding, leading to abnormal microglial coverage around the plaques and increased tau hyperphosphorylation and axonal dystrophy around plaques(*27*, *28*).

Unsurprisingly, the mounting of the microglial response at the transcriptional level, which has been studied most extensively, is significantly reduced in mouse models of AD lacking Trem2 or mice with a knocked-in human R47H variant (*25*, *29*, *30*). Although microglia transcriptional response to AD pathology is largely distinct from mouse models, its dependence on TREM2 is likely, with studies showing that brains of R47H carriers show an altered microglial activation state and induced pluripotent derived microglia carrying this mutation showed a reduced signaling in response to exogenous TREM2 ligands(*31–33*). In addition to TREM2, several other AD risk genes, including PLCG2 and APOE, the strongest risk factor for AD, were shown to alter microglial activation states, strongly suggesting that microglial activation, at least in some contexts or stages of disease, could reflect a protective mechanism in the brain(*34*, *35*). Given that a similar transcriptional signature is observed in other mouse models of neurodegeneration, it is possible that this response could be established in an attempt to deal with neurons enduring an insult, such as being localized around amyloid plaques, containing protein aggregates, undergoing myelin degradation and others(*25*). Hence, we can hypothesize that the induction of putative microglial proteins in the CSF of Lauriet study participants upon treatment with semorinemab and the similarity of that response to the activated state already present in AD brain could indicate an augmentation of the protective microglial activation state, established in response to pathology. Consequently, this may suggest that the distinct nature of the proteomic signatures in Lauriet and Tauriel studies could be reflective of the varying degrees of microglial activation at different degrees of pathology burden in the brain.

One of the limitations of this study is the relatively modest size of our cohort, which potentially leads to a lack of power in detecting differentially abundant proteins as well as associations between proteins and clinical traits, especially when stratifying into smaller patient groups. To this end, the log2 fold changes we report for both Tauriel and Lauriet trial signatures are relatively modest. Further generation of larger CSF proteomics datasets will allow the field to better understand the molecular responses to anti-tau therapies. Additionally, in this study, orthogonal snRNA-seq datasets were leveraged to map proteins to their putative cell types of origins. These results should be interpreted with caution since the correlations between mRNA levels in the brain and protein expression in the CSF can vary. With further development of single-cell proteomics datasets, however, mapping of the brain proteome to the CSF proteome in a cell-type specific manner would become more feasible.

Overall, in this study, we generate a large CSF proteomics dataset that characterizes the pharmacodynamic response of semorinemab and augments the clinical perspective with molecular insights, contributing to the ongoing discourse on tau-targeted candidate therapies and their potential in the complex landscape of AD.

## Materials and Methods

### Sample preparation

CSF samples were shipped frozen and proteins were reducted, alkylated, and digested into peptides using trypsin (Promega, 1:50 protease to total protein ratio) per sample overnight at 37 °C. Peptides were desalted using a HLB μElution plate (Waters) according to the manufacturer’s instructions and dried down using a SpeedVac system. Peptides were resuspended in 1 % acetonitrile and 0.1 % formic acid and spiked with Biognosys’ iRT kit calibration peptides. Peptide concentrations were determined using a UV/VIS Spectrometer at 280 nm (SPECTROstar Nano, BMG Labtech).

### Mass Spectrometry Data Acquisition

For spectral library generation, a sample pool was generated from aliquots of each sample. High-pH reversed-phase chromatography (HPRP) was used to fractionate the pooled sample. Ammonium hydroxide was added to a pH value > 10. The fractionation was performed using a Dionex UltiMate 3000 RS pump (Thermo Scientific) on an Acquity UPLC CSH C18 1.7 μm, 2.1x150 mm column (Waters). The gradient was 1% to 40% solvent B in 30 min, solvents were A: 20 mM ammonium formate in water, B: acetonitrile. Fractions were taken every 30 seconds and sequentially pooled to 16 fraction pools. These were dried down and resuspended in solvent A. Prior to mass spectrometric analyses, they were spiked with Biognosys’ iRT kit calibration peptides.

Peptide concentrations were determined using a UV/VIS Spectrometer at 280nm (SPECTROstar Nano, BMG Labtech). Shotgun DDA LC-MS/MS of HPRP fractions was then performed. Briefly, 3.5 μg of peptides per sample were injected to an in-house packed reversed phase column on a Thermo ScientificTM NeoVanquish UHPLC connected to a Thermo ScientificTM OrbitrapTM Exploris 480TM mass spectrometer equipped with a Nanospray FlexTM ion source and a FAIMS ProTM ion mobility device (Thermo ScientificTM). LC solvents were A: water with 0.1 % FA; B: 80 % acetonitrile, 0.1 % FA in water. The nonlinear LC gradient was 1 – 50 % solvent B in 210 minutes followed by a column washing step in 90 % B for 10 minutes, and a final equilibration step of 1 % B for 8 minutes at 60 °C with a flow rate set to 250 nL/min. MS1 precursor scans were acquired between 330-1650 m/z at 60,000 resolution with data-dependent MS2 scans at 15,000 resolution. MS2 scans were acquired in cycle time of between 1.8 and 2 s per applied FAIMS compensation voltage. Only precursors with charge state 2-6 were isolated for dependent MS2 scans.

For analysis of individual samples, a DIA LC-MS/MS method was used. Samples were reduced, alkylated, and digested with trypsin, and resultant peptides were separated using the same chromatography and MS system described above. The acquisition method used was a FAIMS-DIA method that consisted of one full range MS1 scan and 34 DIA segments as adopted from Bruderer et al. and Tognetti et al. per duty cycle(*36*, *37*).

### Mass Spectrometry Data Analysis

The mass spectrometric DDA data on the pooled CSF HPRP fractions for the spectral library were analyzed using the Pulsar search engine in the Spectronaut software (version 16.2), the false discovery rate on peptide and protein level was set to 1% to generate a library for use in the DIA data analysis. A human UniProt .fasta database (Homo sapiens, 2022-07-01) was used for the search engine, allowing for 2 missed cleavages and variable modifications (N-term acetylation, methionine oxidation, deamidation (NQ) and ammonia-loss). For final data analysis the spectral library created from DDA data was used.

Mass spectrometric data from individual samples collected with FAIMS-DIA were processed using the Spectronaut software (Biognosys, version 16.2), applying the spectral library generated as described earlier. The false discovery rate was set to 1% at both peptide and protein levels, data was filtered using row-based extraction.

Measurements analyzed with Spectronaut were subjected to global median normalization. Feature-level data was exported from Spectronaut and was further analyzed using the ‘MSstats’ package in R to derive protein abundances. All proteins had to be described by at least two unique precursors across all injections. Peptides were filtered on 0.01 Q value, and protein abundances were generated using the ‘dataProcess’ function in ‘MSStats’ by selecting the top 30 features per protein and performing a Tukey Median Polish to assign protein abundances. Further filtering required all samples to identify at least 50% of the total detected proteins in the dataset. For WGCNA and trial proteome signatures, further protein filtering was performed and proteins had to be identified in at least 20% of samples.

### WGCNA network construction

We constructed a weighted protein coexpression network of the batch-corrected CSF proteomics dataset using the Weighted Correlation Network Analysis (WGCNA 1.66) R package. The settings applied were as follows: we set a soft threshold power beta = 17 to transform the adjacency into a signed Topological Overlap Matrix (TOM) using ’min’ as the denominator. Using the TOM-derived dissimilarity, we performed hierarchical clustering. Modules were then identified using a dynamic tree cut method with a deepSplit = 4, respecting the structure of the dendrogram. We applied a minimum module size = 10 to denote the smallest allowable number of proteins in each module. Highly similar modules were subsequently merged at a cut height = 0.12. Numeric labels were assigned to each of the resulting modules. Following the merging, proteins in each module were sorted based on their module membership values (kME scores). Correlation of the modules with clinical scoring metrics was achieved using the Spearman correlation method, with the calculated significance based on non-adjusted p-values.

Samples were run on two separate instruments and an instrument batch effect was observed (Supplemental Figure 8). In order to avoid clustering based on technical variance, a batch-corrected dataset was generated exclusively for the WGCNA analysis. Importantly, because treatment groups and study samples were intentionally balanced across the two instruments, the non-batch corrected dataset was used for all other analyses. For WGCNA, the batch correction was performed on protein-level data using the ComBat method in the *sva* package in R to minimize the instrument effect.

### Differential protein abundance analysis

Baseline and follow-up samples from each patient were processed on the same instrument to mitigate potential instrument batch effects. Differential protein abundance analysis was performed on non-batch corrected and unfiltered data. The “limma” package in R was used to draw comparisons between baseline and follow-up time points within the same patient for each trial and treatment arm.

Trial arm signatures were defined by selecting proteins that are significantly more abundant (FDR < 0.1, LFC > 0) in follow-up relative to baseline in the treatment arms and were detected in at least 80% of samples (Figure 5 A-B). Only the 4500 mg arm data was used. IgG proteins were manually removed. Notably, no proteins were differentially abundant in the placebo arms in either trial.

We evaluated the significance of the Lariet trial signature by permutation testing (Supplementary Figure 6). In each permutation, we randomly selected a gene set with the same length and distribution as the Lariet trial signature from all the detected proteins and recorded the delta between the gene set score of the random list between microglia and the mean of all other cell types.

### Processing of individual sc/snRNA-seq datasets

Three human snRNA-seq datasets derived from the human brain and the CSF were collected from public repositories and re-analyzed. In the case of Morabito et al. and SEA-AD, raw datasets were analyzed with an in-house pipeline. Briefly, reads were demultiplexed based on perfect matches to the expected cell barcodes. Transcript reads were aligned to the human reference genome (GRCh38) using GSNAP(*38*). Only uniquely mapping reads were considered for downstream analysis. Transcript counts for a given gene were based on the number of unique UMIs (up to one mismatch) for reads overlapping exons in sense orientation. Cell barcodes from empty droplets were filtered by requiring a minimum number of detected transcripts. Sample quality was further assessed based on the distribution of per-cell statistics, such as total number of reads, percentage of reads mapping uniquely to the reference genome, percentage of mapped reads overlapping exons, number of detected transcripts (UMIs), number of detected genes, and percentage of mitochondrial transcripts. In the case of Morabito et al., we identified and removed likely ambient RNA contamination with CellBender(*15*, *39*).

Briefly, we ran CellBender to estimate the fraction of ambient RNA from filtered CellRanger counts. In the case of Gate et al., the author-provided count matrix was directly used for the analysis. For all three datasets, author-provided cell type labels were used for downstream analysis.

### Pseudo-bulk analysis of sc/snRNA-seq datasets

Pseudobulk profiles were derived from the SEA-AD, Morabito et al., and Gate et al. datasets by aggregating counts for every cell type (*n*) within each sample (*m*), resulting in *n* X *m* pseudobulks, as previously described(*18*). Pseudobulks containing less than 20 cells were discarded. For each pseudobulk profile, raw counts were generated by adding the total number of UMIs for each gene across all the cells belonging to a particular sample and cell type using the *aggregateAcrossCells* function from the *scuttle* package. This resulted in a gene-by-pseudobulk count matrix which was normalized using the CPM approach from edgeR (with default parameters, except that *prior.count* parameter was set to 3)(*40*). These normalized pseudobulk profiles were then used for differential gene expression analysis between groups of interest and for calculation and visualization of gene set scores.

### Cell type marker identification in the SEA-AD dataset

For the generation of the cell-type markers associated, pseudobulk profiles were derived from the SEA-AD dataset (as described above) and as previously described(*18*). Briefly, expression profiles of every cell type were compared to all other cell types, resulting in *n - 1* contrasts per cell type. Significant genes were selected by filtering for FDR < 0.001 and LFC > 3 for every cell type-to-cell type comparison, resulting in *n - 1* marker gene lists. Final markers were selected as those genes that had an LFC of at least 1 in at least 5 contrasts. We used Vip (vasoactive intestinal peptide-expressing interneurons) neurons as a representative population for inhibitory neurons and Layer 2/3 neurons as representative for excitatory neurons since they were the most abundant neuronal subtypes within their respective categories. All differential expression analysis was performed with *voom*(*41*).

### Module cell-type enrichment analysis

Enrichment analysis was performed using a hypergeometric test, specifically the ‘phyper’ function from the ‘stats’ package in R, on the sets of proteins found within each module and sets of cell-type specific markers derived from SEA-AD dataset to ascertain the cell-type enrichments present. Non-adjusted p-values were used to evaluate the enrichment significance.

### Gene set analysis

Gene/protein set scores (Figures 6, S3, S5, S6, S7) were calculated as previously described(*42*). Briefly, gene expression values from snRNA-seq datasets were first log-transformed and stabilized as Log2(nCPM+1).The gene set score (based on gene expression) or protein set score (based on protein abundance derived from MSstats) for a sample was then calculated as the average over all genes or proteins within a gene/protein set.

The workflow figure (Figure 1) was generated using BioRender.com(*43*).

## Data Availability

Patient-level data will not be made available upon publication.

---

**Supplementary Figure 1.**
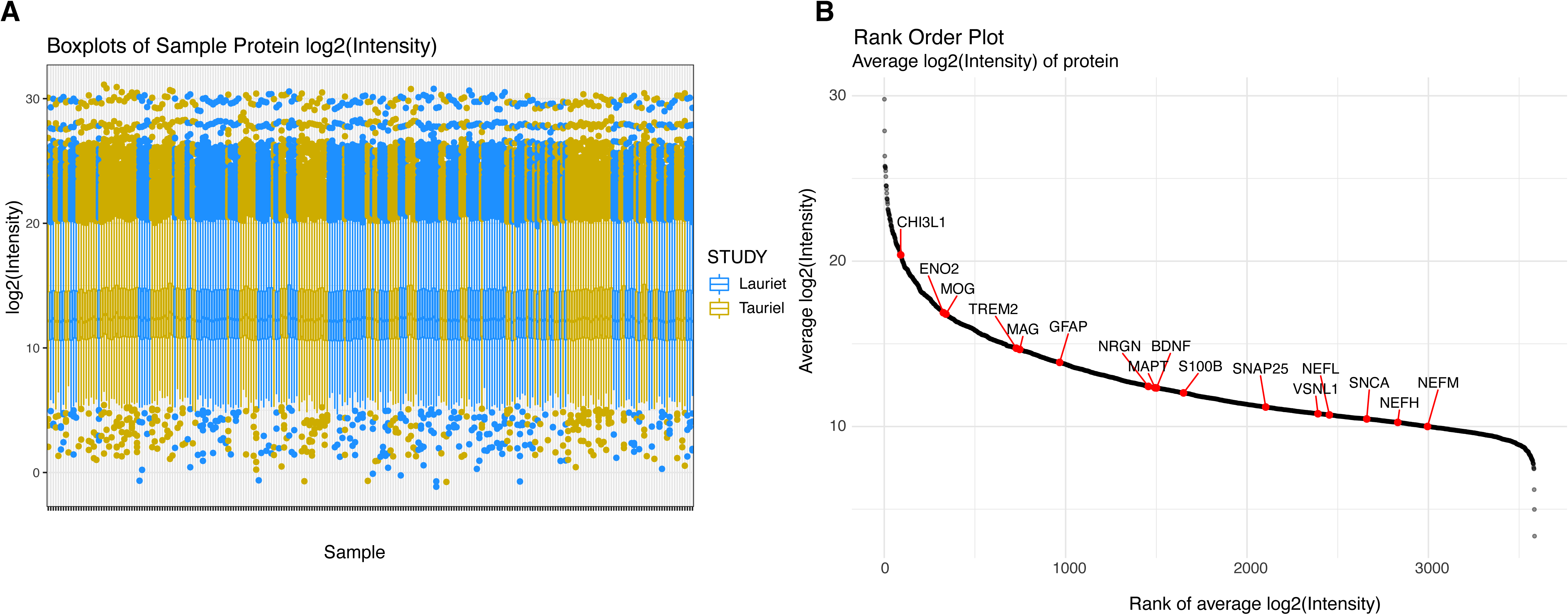
Dataset background metrics. **A.** Boxplots showing median normalized log2 protein intensities for all samples in the dataset, color-coded by their respective study. Protein intensities were acquired through DIA-MS on non-depleted CSF using 3.8 hour gradients. **B.** Ranking of unique proteins in the dataset based on average log2 protein intensities across all samples in the dataset. Select proteins of interest are highlighted.

**Supplementary Figure 2.**
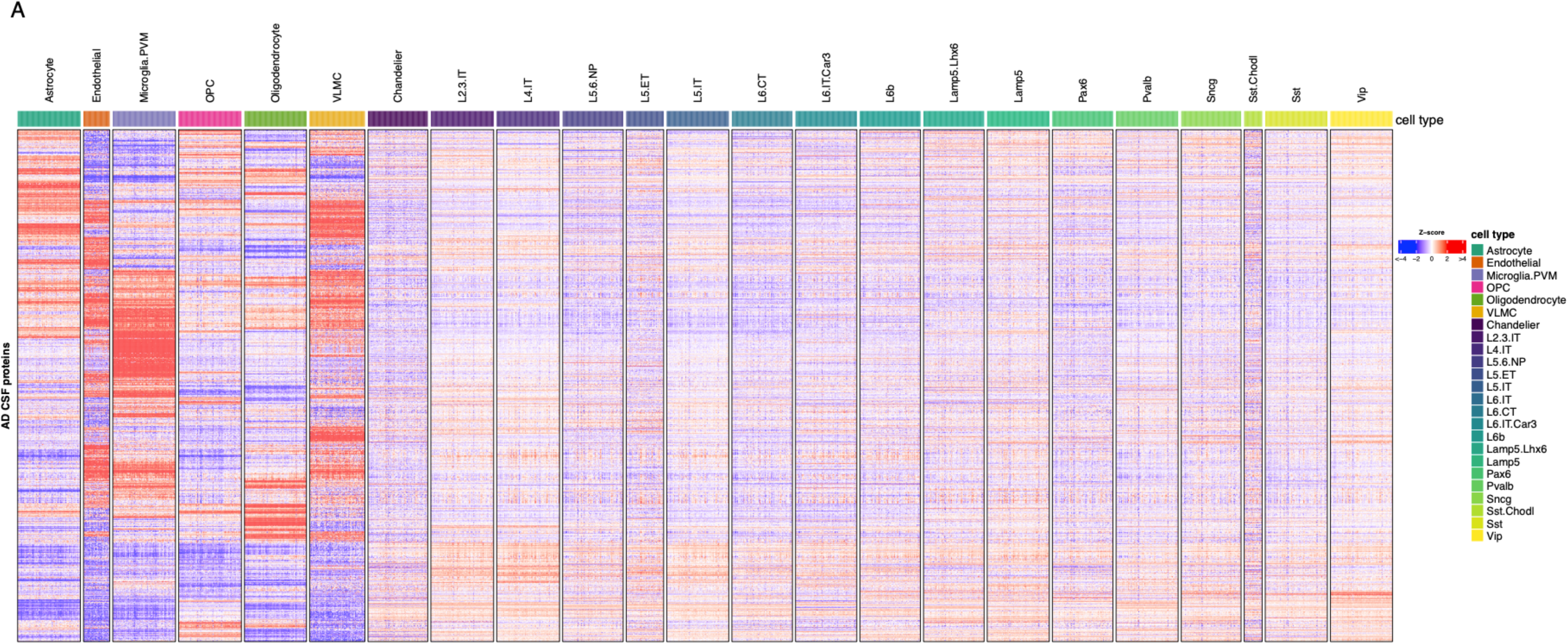
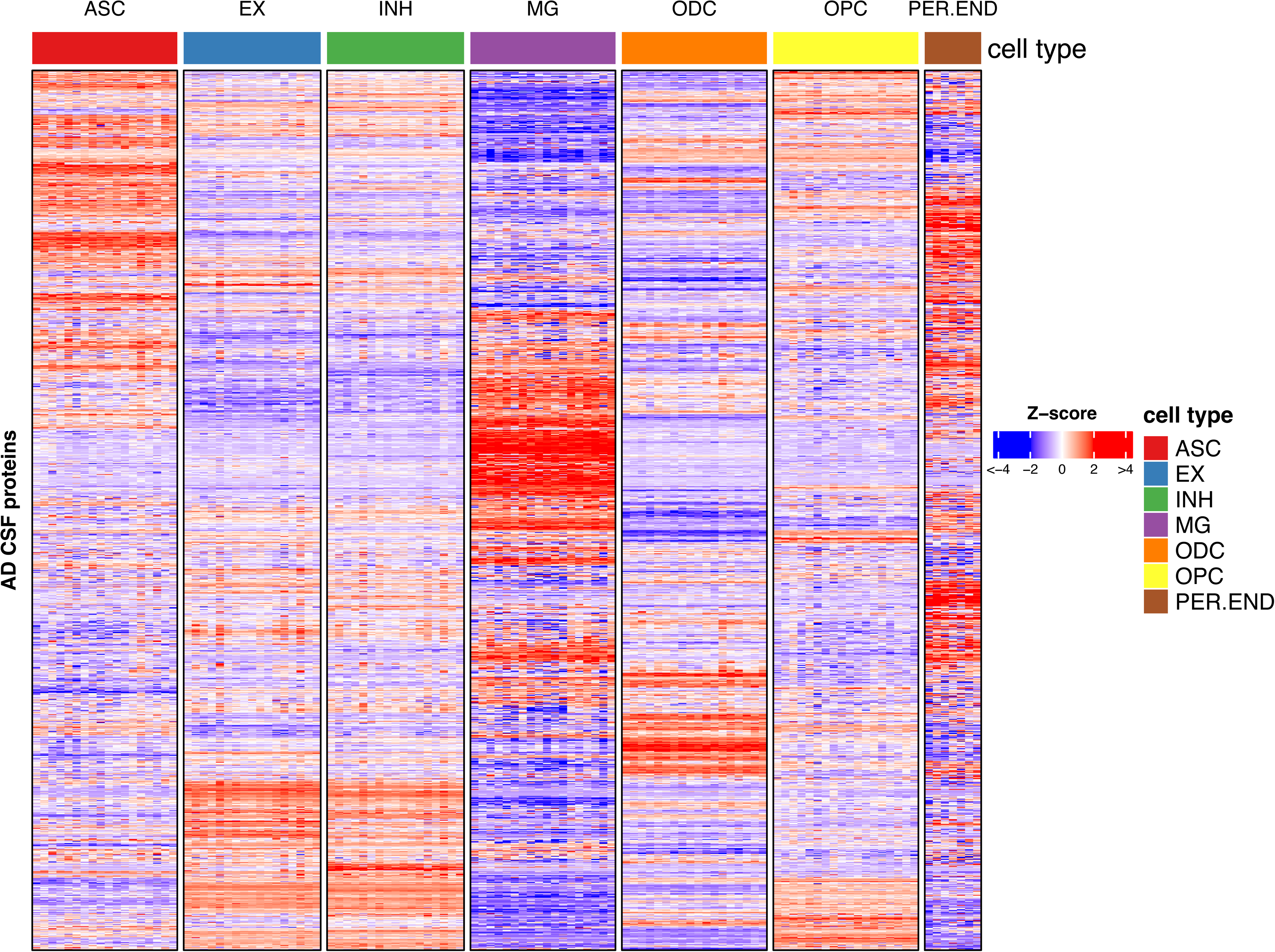
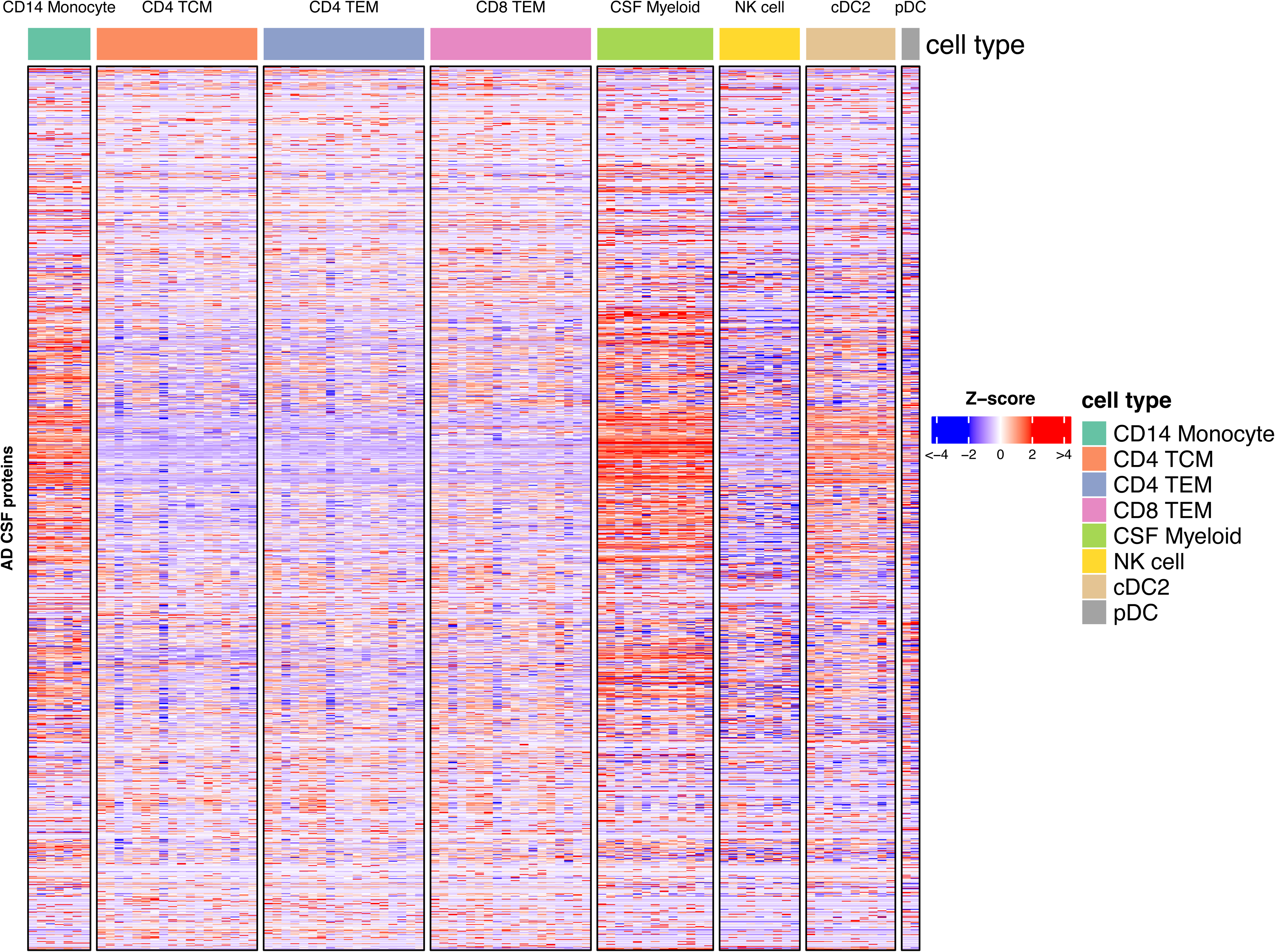
Expression of genes encoding AD CSF proteins across the brain and CSF resident cell types. Heatmap showing row-normalized *Z* scores for genes encoding proteins detected in the CSF across pseudobulk replicates from **A)** all cell types from the brain snRNA-seq SEA-AD dataset (including all excitatory and inhibitory neuronal subtypes), **B)** cell types from brain snRNA-seq Morabito et al., dataset, and **C)** Gate et al., CSF snRNA-seq dataset. The row order of genes is the same in A-C. Each row represents the normalized gene expression level of the encoded protein. Each column is an individual cell-type specific donor pseudobulk.

**Supplementary Figure 3.**
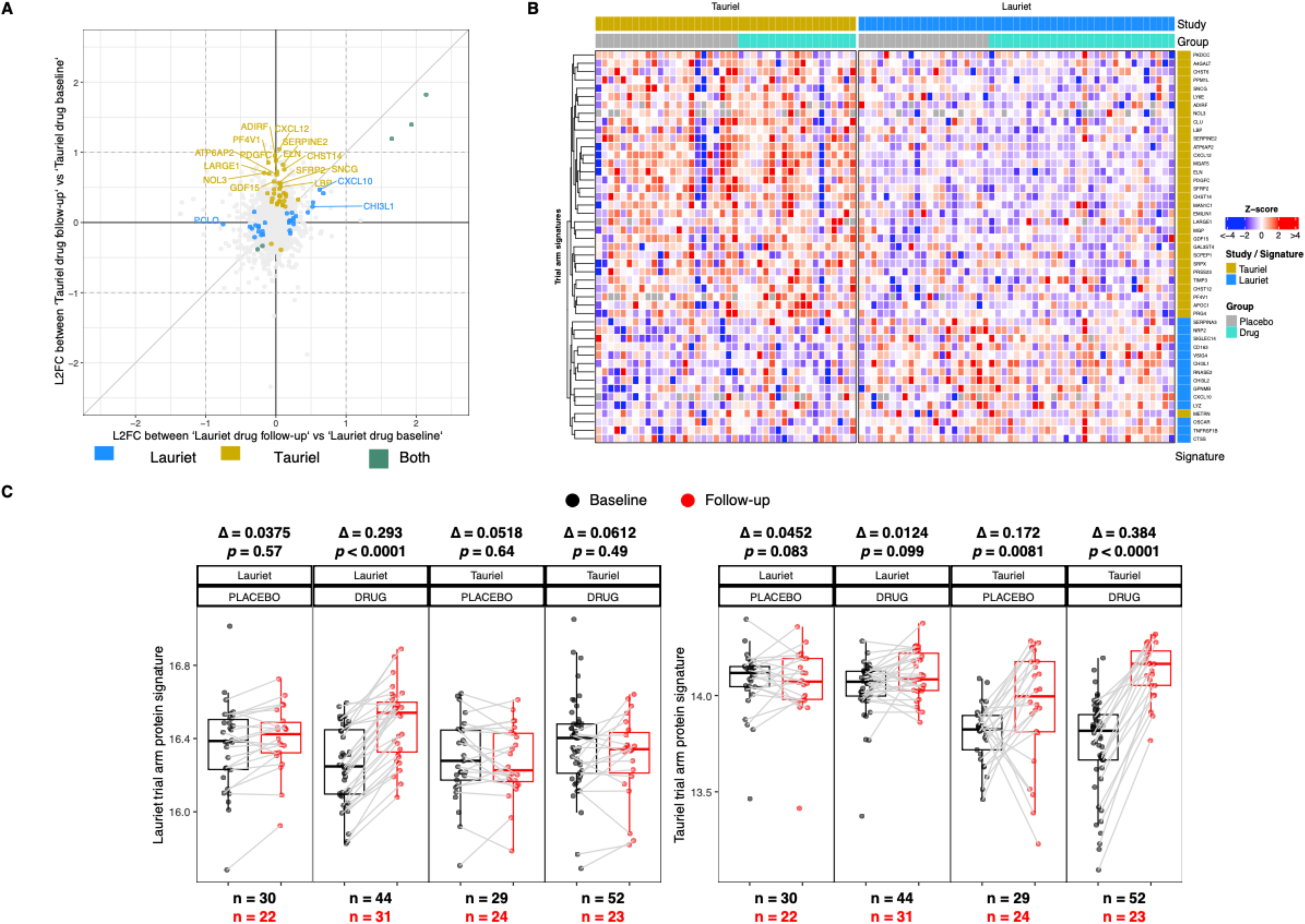
Lauriet and Tauriel protein signatures are distinct. **A.** 4-way comparison of log fold changes in Lauriet (x-axis) and Tauriel (y-axis) trial arm protein signatures prior to filtering. Each point represents one protein colored by whether FDR was < 0.1 in one or both trials (golden for Tauriel, blue for Lauriet and cyan for both). **B.** Heatmap showing row-normalized Z scores for Lauriet and Tauriel signature proteins based on log2 fold change between follow-up relative to baseline samples in each group. **C.** Distribution of protein set scores for Lauriet and Tauriel trial arm protein signatures. *Δ*, mean log2 fold change; *p-value*, Wilcoxon test.

**Supplementary Figure 4.**
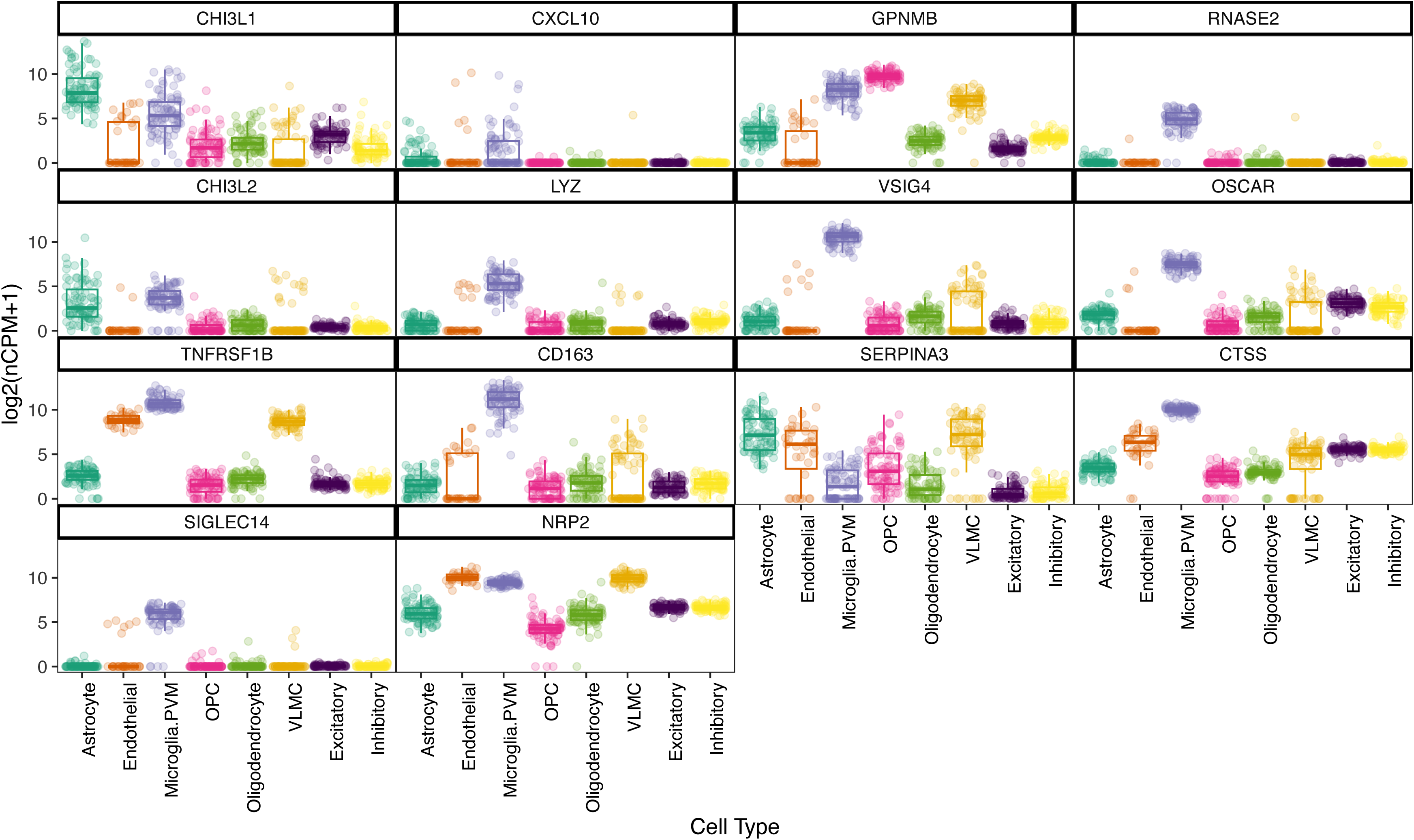
Individual genes encoding proteins in the Lauriet trial arm signature are largely enriched in microglia. Distribution of gene set scores for individual genes encoding proteins in the Lauriet trial arm signature across pseudo bulk profiles of cell types from the SEA-AD brain dataset.

**Supplementary Figure 5.**
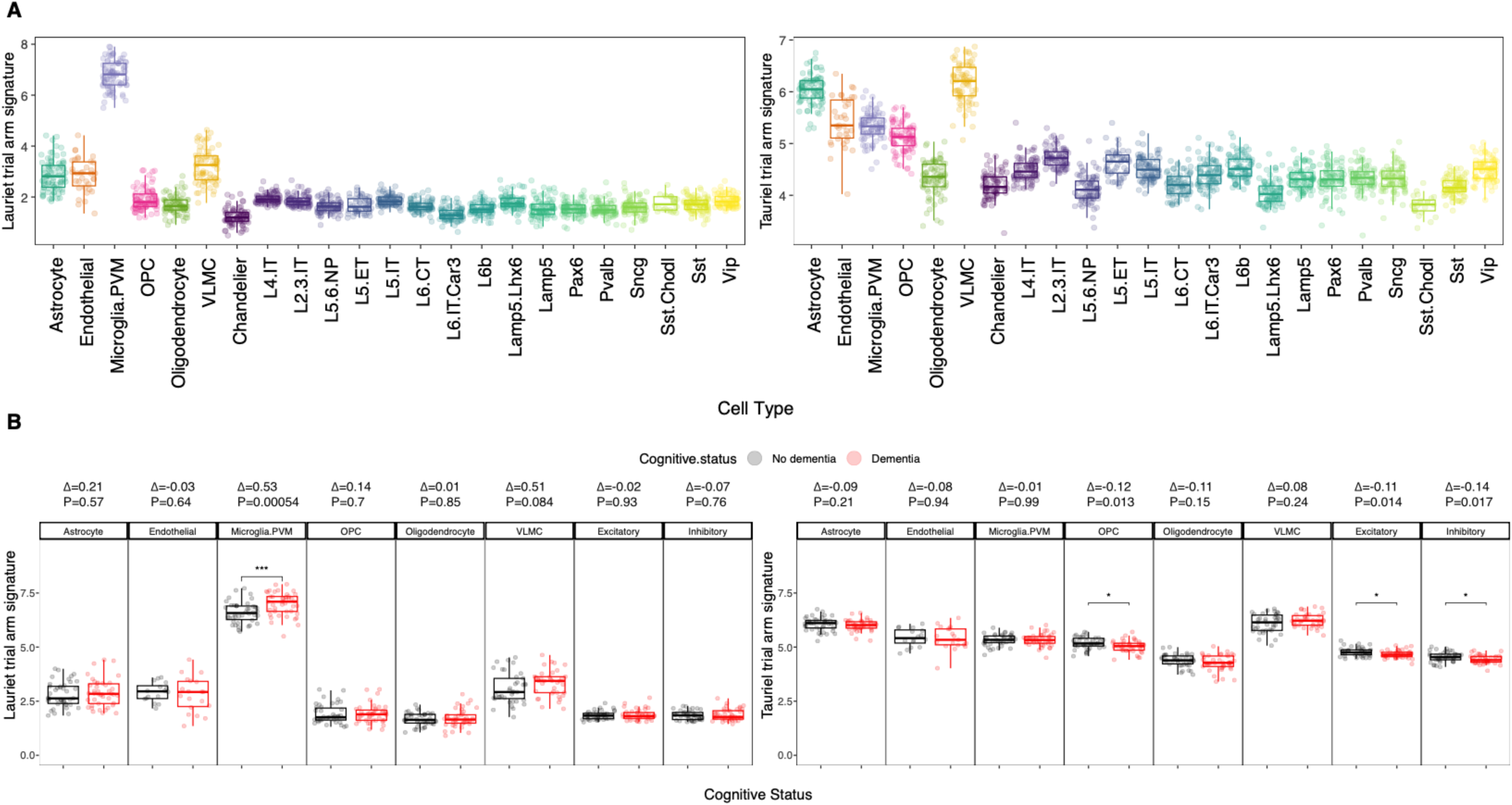
Lauriet trial gene signature is enriched in microglial cells, while Lauriet trial gene signature is expressed more broadly. Distribution of Lauriet and Tauriel gene signatures across pseudobulk profiles in **A)** all cell types from the SEA-AD brain dataset and **B)** comparing demented *vs.* non-demented subjects across cell types. *Δ*, mean log2 fold change; *p-value*, Wilcoxon test. Excitatory and Inhibitory labels refer to Vip (vasoactive intestinal peptide-expressing interneurons) and L2/3 neuronal layer as the most abundant excitatory and inhibitory neuronal subtypes in the SEA-AD dataset.

**Supplementary Figure 6.**
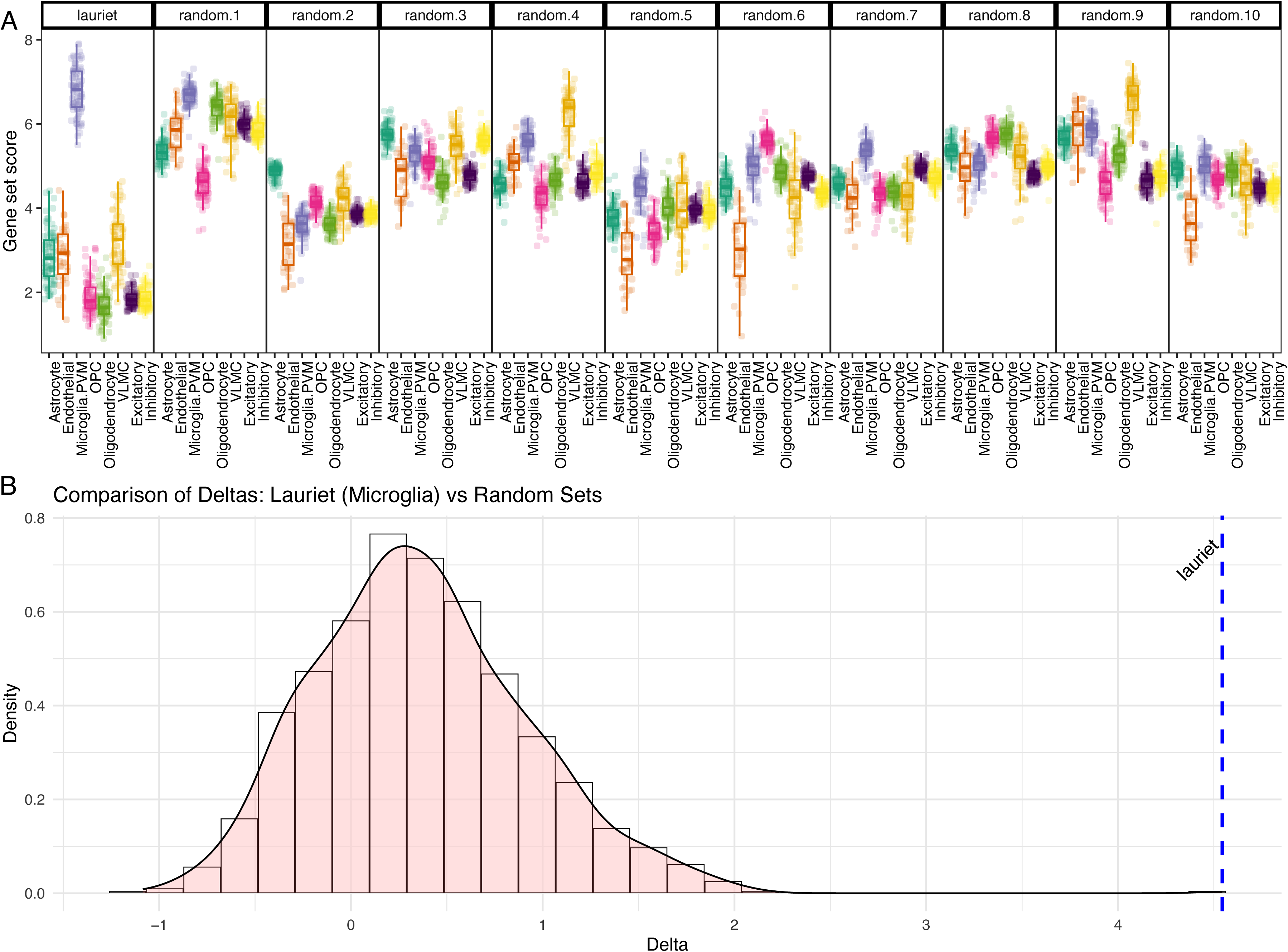
Permutational testing to assess significance of Lauriet signature enrichment in microglial cells. For each permutation test, a gene set was randomly selected from detected proteins (filtered for missingness) such that the overall distribution of the random gene set overlaps that of the Lauriet trial arm gene set. Delta was then computed between the gene set score of the random set in pseudobulk profile of microglia cell type from SEA-AD relative to the mean of the gene set score in pseudobulk profiles of all other cell types. **A)** Representative 10 permutation tests showing that the gene set score of Lauriet trial arm signature is significantly enriched in microglia relative to all other cell types while randomly selected gene sets are not enriched in microglia. **B)** Distribution of deltas between microglia gene set score of a random protein set and mean score of all other cell types based on 100 permutations.

**Supplementary Figure 7.**
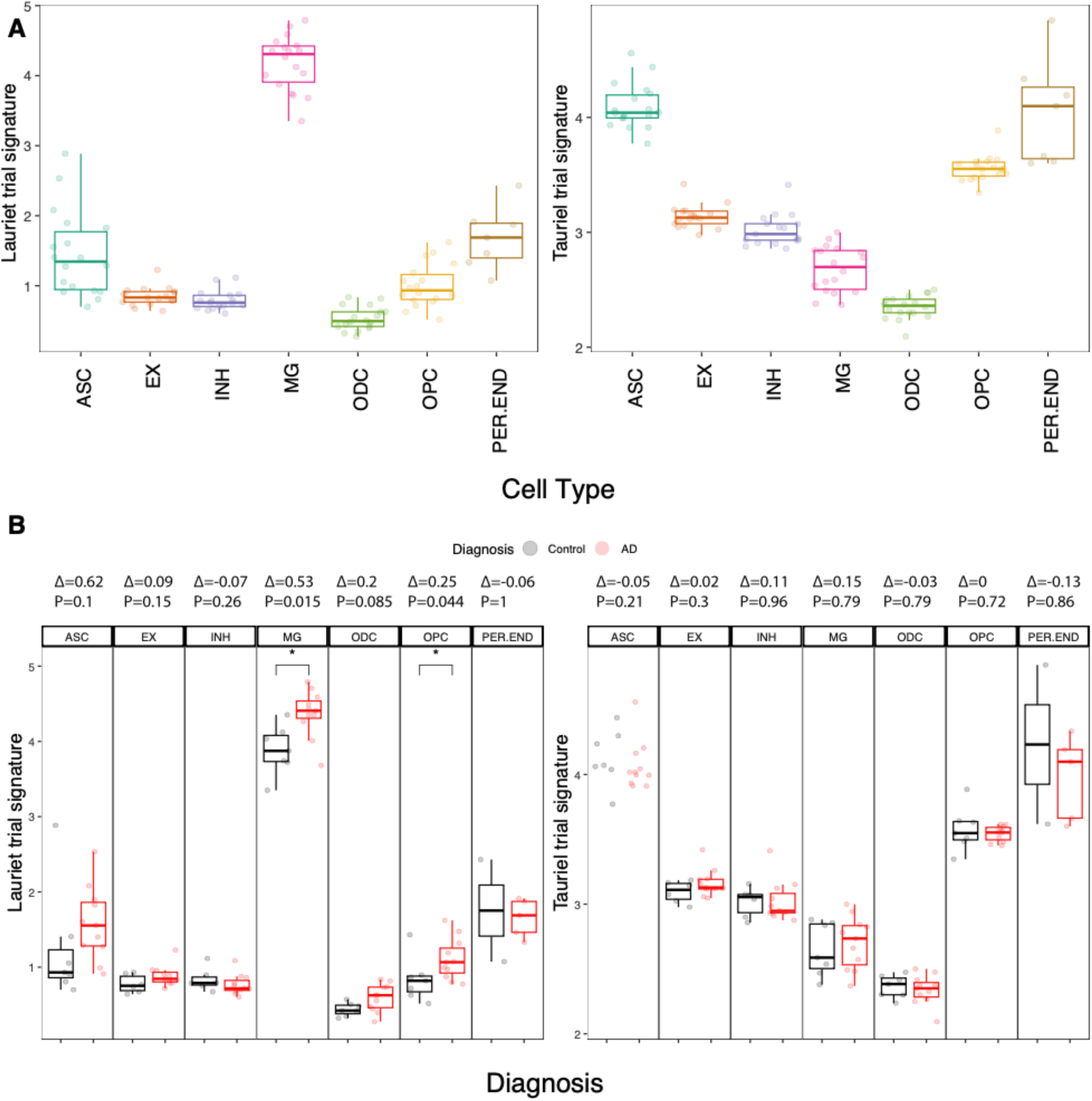
Validation of the microglial enrichment of the Lauriet gene signature in Morabito *et al.*, brain snRNA-seq dataset. Distribution of Lauriet and Tauriel gene signatures across pseudobulk profiles in **A)** cell types from the Morabito et al. and **B)** comparing AD *vs.* control subjects in the same set of cell types. *Δ*, mean log2 fold change; *p-value*, Wilcoxon test.

**Supplementary Figure 8.**
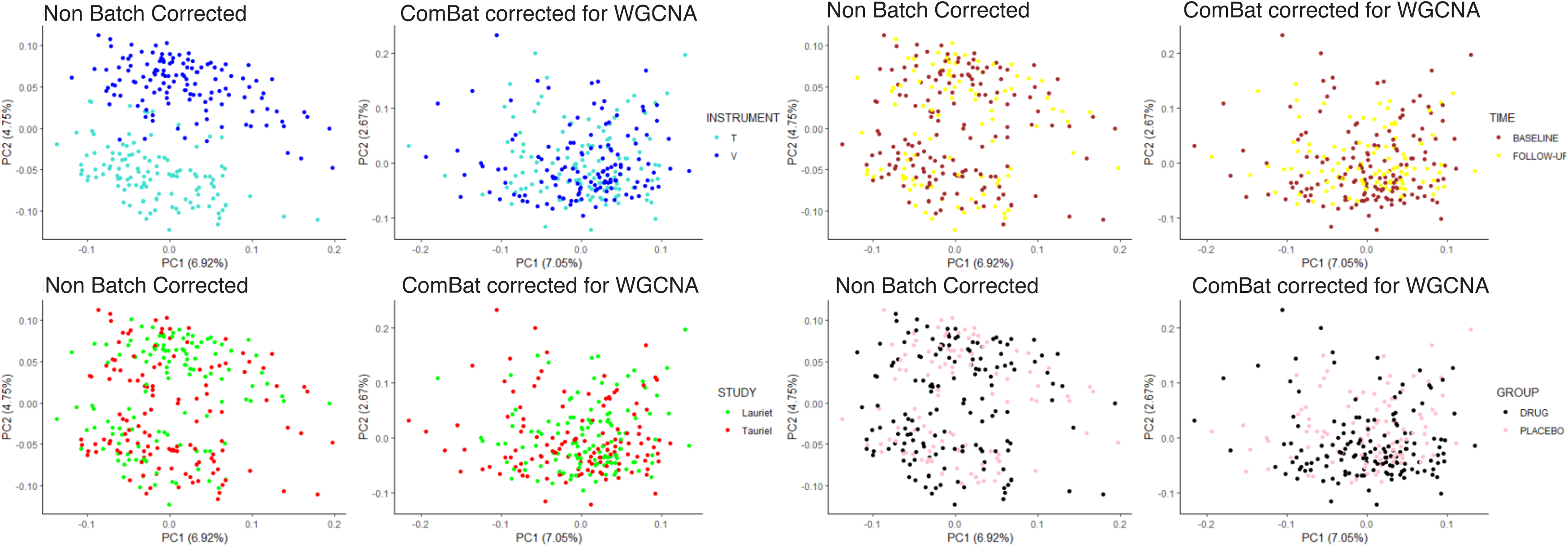
Pre-post batch correction PCAs. PCAs were generated to investigate variables influencing sample separation before and after instrument batch correction. Plots A-D depict the influence and removal of different variables including Instrument (T or V), Time (Baseline or Follow up), Study (Tauriel or Lauriet), and Group (Drug or Placebo).

## References

1. D. S. Knopman, H. Amieva, R. C. Petersen, G. Chételat, D. M. Holtzman, B. T. Hyman, R. A. Nixon, D. T. Jones, Alzheimer disease. Nature Reviews Disease Primers 7, 1–21 (2021).

2. P. T. Nelson, I. Alafuzoff, E. H. Bigio, C. Bouras, H. Braak, N. J. Cairns, R. J. Castellani, B. J. Crain, P. Davies, K. Del Tredici, C. Duyckaerts, M. P. Frosch, V. Haroutunian, P. R. Hof, C. M. Hulette, B. T. Hyman, T. Iwatsubo, K. A. Jellinger, G. A. Jicha, E. Kövari, W. A. Kukull, J. B. Leverenz, S. Love, I. R. Mackenzie, D. M. Mann, E. Masliah, A. C. McKee, T. J. Montine, J. C. Morris, J. A. Schneider, J. A. Sonnen, D. R. Thal, J. Q. Trojanowski, J. C. Troncoso, T. Wisniewski, R. L. Woltjer, T. G. Beach, Correlation of Alzheimer disease neuropathologic changes with cognitive status: a review of the literature. J. Neuropathol. Exp. Neurol. 71, 362–381 (2012).

3. E. Teng, P. T. Manser, K. Pickthorn, F. Brunstein, M. Blendstrup, S. Sanabria Bohorquez, K. R. Wildsmith, B. Toth, M. Dolton, V. Ramakrishnan, A. Bobbala, S. A. M. Sikkes, M. Ward, R. N. Fuji, G. A. Kerchner, Tauriel Investigators, Safety and Efficacy of Semorinemab in Individuals With Prodromal to Mild Alzheimer Disease: A Randomized Clinical Trial. JAMA Neurol. 79, 758–767 (2022).

4. G. Ayalon, S.-H. Lee, O. Adolfsson, C. Foo-Atkins, J. K. Atwal, M. Blendstrup, H. Booler, J. Bravo, R. Brendza, F. Brunstein, R. Chan, P. Chandra, J. A. Couch, A. Datwani, B. Demeule, D. DiCara, R. Erickson, J. A. Ernst, O. Foreman, D. He, I. Hötzel, M. Keeley, M. C. M. Kwok, J. Lafrance-Vanasse, H. Lin, Y. Lu, W. Luk, P. Manser, A. Muhs, H. Ngu, A. Pfeifer, M. Pihlgren, G. K. Rao, K. Scearce-Levie, S. P. Schauer, W. B. Smith, H. Solanoy, E. Teng, K. R. Wildsmith, D. B. Yadav, Y. Ying, R. N. Fuji, G. A. Kerchner, Antibody semorinemab reduces tau pathology in a transgenic mouse model and engages tau in patients with Alzheimer’s disease. Sci. Transl. Med., doi: 10.1126/scitranslmed.abb2639 (2021).

5. C. Monteiro, B. Toth, F. Brunstein, A. Bobbala, S. Datta, R. Ceniceros, S. M. Sanabria Bohorquez, V. G. Anania, K. R. Wildsmith, S. P. Schauer, J. Lee, M. J. Dolton, V. Ramakrishnan, D. Abramzon, E. Teng, Lauriet investigators, Randomized Phase II Study of the Safety and Efficacy of Semorinemab in Participants With Mild-to-Moderate Alzheimer Disease: Lauriet. Neurology 101, e1391–e1401 (2023).

6. N. T. Seyfried, E. B. Dammer, V. Swarup, D. Nandakumar, D. M. Duong, L. Yin, Q. Deng, T. Nguyen, C. M. Hales, T. Wingo, J. Glass, M. Gearing, M. Thambisetty, J. C. Troncoso, D. H. Geschwind, J. J. Lah, A. I. Levey, A Multi-network Approach Identifies Protein-Specific Co-expression in Asymptomatic and Symptomatic Alzheimer’s Disease. Cell Syst 4, 60–72.e4 (2017).

7. L. Higginbotham, E. B. Dammer, D. M. Duong, E. Modeste, T. J. Montine, J. J. Lah, A. I. Levey, N. T. Seyfried, Network Analysis of a Membrane-Enriched Brain Proteome across Stages of Alzheimer’s Disease. Proteomes 7 (2019).

8. L. Higginbotham, L. Ping, E. B. Dammer, D. M. Duong, M. Zhou, M. Gearing, C. Hurst, J. D. Glass, S. A. Factor, E. C. B. Johnson, I. Hajjar, J. J. Lah, A. I. Levey, N. T. Seyfried, Integrated proteomics reveals brain-based cerebrospinal fluid biomarkers in asymptomatic and symptomatic Alzheimer’s disease. Sci Adv 6 (2020).

9. B. M. Tijms, E. M. Vromen, O. Mjaavatten, H. Holstege, L. M. Reus, S. van der Lee, K. E. J. Wesenhagen, L. Lorenzini, L. Vermunt, V. Venkatraghavan, N. Tesi, J. Tomassen, A. den Braber, J. Goossens, E. Vanmechelen, F. Barkhof, Y. A. L. Pijnenburg, W. M. van der Flier, C. E. Teunissen, F. S. Berven, P. J. Visser, Cerebrospinal fluid proteomics in patients with Alzheimer’s disease reveals five molecular subtypes with distinct genetic risk profiles. Nat Aging 4, 33–47 (2024).

10. Applied Research Applied Research Press, WGCNA: An R Package for Weighted Correlation Network Analysis (2015).

11. S. Sanabria Bohórquez, J. Marik, A. Ogasawara, J. N. Tinianow, H. S. Gill, O. Barret, G. Tamagnan, D. Alagille, G. Ayalon, P. Manser, T. Bengtsson, M. Ward, S.-P. Williams, G. A. Kerchner, J. P. Seibyl, K. Marek, R. M. Weimer, [F]GTP1 (Genentech Tau Probe 1), a radioligand for detecting neurofibrillary tangle tau pathology in Alzheimer’s disease. Eur. J. Nucl. Med. Mol. Imaging 46, 2077–2089 (2019).

12. A. P. Wingo, E. B. Dammer, M. S. Breen, B. A. Logsdon, D. M. Duong, J. C. Troncosco, M. Thambisetty, T. G. Beach, G. E. Serrano, E. M. Reiman, R. J. Caselli, J. J. Lah, N. T. Seyfried, A. I. Levey, T. S. Wingo, Large-scale proteomic analysis of human brain identifies proteins associated with cognitive trajectory in advanced age. Nat. Commun. 10, 1619 (2019).

13. M. I. Gabitto, K. J. Travaglini, V. M. Rachleff, E. S. Kaplan, B. Long, J. Ariza, Y. Ding, J. T. Mahoney, N. Dee, J. Goldy, E. J. Melief, K. Brouner, J. Campos, A. J. Carr, T. Casper, R. Chakrabarty, M. Clark, J. Compos, J. Cool, N. J. Valera Cuevas, R. Dalley, M. Darvas, S.-L. Ding, T. Dolbeare, C. L. Mac Donald, T. Egdorf, L. Esposito, R. Ferrer, R. Gala, A. Gary, J. Gloe, N. Guilford, J. Guzman, W. Ho, T. Jarksy, N. Johansen, B. E. Kalmbach, L. M. Keene, S. Khawand, M. Kilgore, A. Kirkland, M. Kunst, B. R. Lee, J. Malone, Z. Maltzer, N. Martin, R. McCue, D. McMillen, E. Meyerdierks, K. P. Meyers, T. Mollenkopf, M. Montine, A. L. Nolan, J. Nyhus, P. A. Olsen, M. Pacleb, T. Pham, C. A. Pom, N. Postupna, A. Ruiz, A. M. Schantz, S. A. Sorensen, B. Staats, M. Sullivan, S. M. Sunkin, C. Thompson, M. Tieu, J. Ting, A. Torkelson, T. Tran, M.-Q. Wang, J. Waters, A. M. Wilson, D. Haynor, N. Gatto, S. Jayadev, S. Mufti, L. Ng, S. Mukherjee, P. K. Crane, C. S. Latimer, B. P. Levi, K. Smith, J. L. Close, J. A. Miller, R. D. Hodge, E. B. Larson, T. J. Grabowski, M. Hawrylycz, C. D. Keene, E. S. Lein, Integrated multimodal cell atlas of Alzheimer’s disease. Res Sq, doi: 10.21203/rs.3.rs-2921860/v1 (2023).

14. A. H. Khasawneh, R. J. Garling, C. A. Harris, Cerebrospinal fluid circulation: What do we know and how do we know it? Brain Circ 4, 14–18 (2018).

15. S. Morabito, E. Miyoshi, N. Michael, S. Shahin, A. C. Martini, E. Head, J. Silva, K. Leavy, M. Perez-Rosendahl, V. Swarup, Single-nucleus chromatin accessibility and transcriptomic characterization of Alzheimer’s disease. Nat. Genet. 53, 1143–1155 (2021).

16. D. Gate, N. Saligrama, O. Leventhal, A. C. Yang, M. S. Unger, J. Middeldorp, K. Chen, B. Lehallier, D. Channappa, M. B. De Los Santos, A. McBride, J. Pluvinage, F. Elahi, G. K.-Y. Tam, Y. Kim, M. Greicius, A. D. Wagner, L. Aigner, D. R. Galasko, M. M. Davis, T. Wyss-Coray, Clonally expanded CD8 T cells patrol the cerebrospinal fluid in Alzheimer’s disease. Nature 577, 399–404 (2020).

17. A. M. Smith, K. Davey, S. Tsartsalis, C. Khozoie, N. Fancy, S. S. Tang, E. Liaptsi, M. Weinert, A. McGarry, R. C. J. Muirhead, S. Gentleman, D. R. Owen, P. M. Matthews, Diverse human astrocyte and microglial transcriptional responses to Alzheimer’s pathology. Acta Neuropathol. 143, 75–91 (2022).

18. S. Pandey, K. Shen, S.-H. Lee, Y.-A. A. Shen, Y. Wang, M. Otero-García, N. Kotova, S. T. Vito, B. I. Laufer, D. F. Newton, M. G. Rezzonico, J. E. Hanson, J. S. Kaminker, C. J. Bohlen, T. J. Yuen, B. A. Friedman, Disease-associated oligodendrocyte responses across neurodegenerative diseases. Cell Rep. 40, 111189 (2022).

19. F. Panza, M. Lozupone, The challenges of anti-tau therapeutics in Alzheimer disease. Nat. Rev. Neurol. 18, 577–578 (2022).

20. J. M. Bader, P. E. Geyer, J. B. Müller, M. T. Strauss, M. Koch, F. Leypoldt, P. Koertvelyessy, D. Bittner, C. G. Schipke, E. I. Incesoy, O. Peters, N. Deigendesch, M. Simons, M. K. Jensen, H. Zetterberg, M. Mann, Proteome profiling in cerebrospinal fluid reveals novel biomarkers of Alzheimer’s disease. Mol. Syst. Biol. 16, e9356 (2020).

21. M. Askenazi, T. Kavanagh, G. Pires, B. Ueberheide, T. Wisniewski, E. Drummond, Compilation of reported protein changes in the brain in Alzheimer’s disease. Nat. Commun. 14, 4466 (2023).

22. M. Prinz, S. Jung, J. Priller, Microglia Biology: One Century of Evolving Concepts. Cell 179, 292–311 (2019).

23. R. Sims, S. J. van der Lee, A. C. Naj, C. Bellenguez, N. Badarinarayan, J. Jakobsdottir, B. W. Kunkle, A. Boland, R. Raybould, J. C. Bis, E. R. Martin, B. Grenier-Boley, S. Heilmann-Heimbach, V. Chouraki, A. B. Kuzma, K. Sleegers, M. Vronskaya, A. Ruiz, R. R. Graham, R. Olaso, P. Hoffmann, M. L. Grove, B. N. Vardarajan, M. Hiltunen, M. M. Nöthen, C. C. White, K. L. Hamilton-Nelson, J. Epelbaum, W. Maier, S.-H. Choi, G. W. Beecham, C. Dulary, S. Herms, A. V. Smith, C. C. Funk, C. Derbois, A. J. Forstner, S. Ahmad, H. Li, D. Bacq, D. Harold, C. L. Satizabal, O. Valladares, A. Squassina, R. Thomas, J. A. Brody, L. Qu, P. Sánchez-Juan, T. Morgan, F. J. Wolters, Y. Zhao, F. S. Garcia, N. Denning, M. Fornage, J. Malamon, M. C. D. Naranjo, E. Majounie, T. H. Mosley, B. Dombroski, D. Wallon, M. K. Lupton, J. Dupuis, P. Whitehead, L. Fratiglioni, C. Medway, X. Jian, S. Mukherjee, L. Keller, K. Brown, H. Lin, L. B. Cantwell, F. Panza, B. McGuinness, S. Moreno-Grau, J. D. Burgess, V. Solfrizzi, P. Proitsi, H. H. Adams, M. Allen, D. Seripa, P. Pastor, L. A. Cupples, N. D. Price, D. Hannequin, A. Frank-García, D. Levy, P. Chakrabarty, P. Caffarra, I. Giegling, A. S. Beiser, V. Giedraitis, H. Hampel, M. E. Garcia, X. Wang, L. Lannfelt, P. Mecocci, G. Eiriksdottir, P. K. Crane, F. Pasquier, V. Boccardi, I. Henández, R. C. Barber, M. Scherer, L. Tarraga, P. M. Adams, M. Leber, Y. Chen, M. S. Albert, S. Riedel-Heller, V. Emilsson, D. Beekly, A. Braae, R. Schmidt, D. Blacker, C. Masullo, H. Schmidt, R. S. Doody, G. Spalletta, W. T. Longstreth Jr, T. J. Fairchild, P. Bossù, O. L. Lopez, M. P. Frosch, E. Sacchinelli, B. Ghetti, Q. Yang, R. M. Huebinger, F. Jessen, S. Li, M. I. Kamboh, J. Morris, O. Sotolongo-Grau, M. J. Katz, C. Corcoran, M. Dunstan, A. Braddel, C. Thomas, A. Meggy, R. Marshall, A. Gerrish, J. Chapman, M. Aguilar, S. Taylor, M. Hill, M. D. Fairén, A. Hodges, B. Vellas, H. Soininen, I. Kloszewska, M. Daniilidou, J. Uphill, Y. Patel, J. T. Hughes, J. Lord, J. Turton, A. M. Hartmann, R. Cecchetti, C. Fenoglio, M. Serpente, M. Arcaro, C. Caltagirone, M. D. Orfei, A. Ciaramella, S. Pichler, M. Mayhaus, W. Gu, A. Lleó, J. Fortea, R. Blesa, I. S. Barber, K. Brookes, C. Cupidi, R. G. Maletta, D. Carrell, S. Sorbi, S. Moebus, M. Urbano, A. Pilotto, J. Kornhuber, P. Bosco, S. Todd, D. Craig, J. Johnston, M. Gill, B. Lawlor, A. Lynch, N. C. Fox, J. Hardy, ARUK Consortium, R. L. Albin, L. G. Apostolova, S. E. Arnold, S. Asthana, C. S. Atwood, C. T. Baldwin, L. L. Barnes, S. Barral, T. G. Beach, J. T. Becker, E. H. Bigio, T. D. Bird, B. F. Boeve, J. D. Bowen, A. Boxer, J. R. Burke, J. M. Burns, J. D. Buxbaum, N. J. Cairns, C. Cao, C. S. Carlson, C. M. Carlsson, R. M. Carney, M. M. Carrasquillo, S. L. Carroll, C. C. Diaz, H. C. Chui, D. G. Clark, D. H. Cribbs, E. A. Crocco, C. DeCarli, M. Dick, R. Duara, D. A. Evans, K. M. Faber, K. B. Fallon, D. W. Fardo, M. R. Farlow, S. Ferris, T. M. Foroud, D. R. Galasko, M. Gearing, D. H. Geschwind, J. R. Gilbert, N. R. Graff-Radford, R. C. Green, J. H. Growdon, R. L. Hamilton, L. E. Harrell, L. S. Honig, M. J. Huentelman, C. M. Hulette, B. T. Hyman, G. P. Jarvik, E. Abner, L.-W. Jin, G. Jun, A. Karydas, J. A. Kaye, R. Kim, N. W. Kowall, J. H. Kramer, F. M. LaFerla, J. J. Lah, J. B. Leverenz, A. I. Levey, G. Li, A. P. Lieberman, K. L. Lunetta, C. G. Lyketsos, D. C. Marson, F. Martiniuk, D. C. Mash, E. Masliah, W. C. McCormick, S. M. McCurry, A. N. McDavid, A. C. McKee, M. Mesulam, B. L. Miller, C. A. Miller, J. W. Miller, J. C. Morris, J. R. Murrell, A. J. Myers, S. O’Bryant, J. M. Olichney, V. S. Pankratz, J. E. Parisi, H. L. Paulson, W. Perry, E. Peskind, A. Pierce, W. W. Poon, H. Potter, J. F. Quinn, A. Raj, M. Raskind, B. Reisberg, C. Reitz, J. M. Ringman, E. D. Roberson, E. Rogaeva, H. J. Rosen, R. N. Rosenberg, M. A. Sager, A. J. Saykin, J. A. Schneider, L. S. Schneider, W. W. Seeley, A. G. Smith, J. A. Sonnen, S. Spina, R. A. Stern, R. H. Swerdlow, R. E. Tanzi, T. A. Thornton-Wells, J. Q. Trojanowski, J. C. Troncoso, V. M. Van Deerlin, L. J. Van Eldik, H. V. Vinters, J. P. Vonsattel, S. Weintraub, K. A. Welsh-Bohmer, K. C. Wilhelmsen, J. Williamson, T. S. Wingo, R. L. Woltjer, C. B. Wright, C.-E. Yu, L. Yu, F. Garzia, F. Golamaully, G. Septier, S. Engelborghs, R. Vandenberghe, P. P. De Deyn, C. M. Fernadez, Y. A. Benito, H. Thonberg, C. Forsell, L. Lilius, A. Kinhult-Stählbom, L. Kilander, R. Brundin, L. Concari, S. Helisalmi, A. M. Koivisto, A. Haapasalo, V. Dermecourt, N. Fievet, O. Hanon, C. Dufouil, A. Brice, K. Ritchie, B. Dubois, J. J. Himali, C. D. Keene, J. Tschanz, A. L. Fitzpatrick, W. A. Kukull, M. Norton, T. Aspelund, E. B. Larson, R. Munger, J. I. Rotter, R. B. Lipton, M. J. Bullido, A. Hofman, T. J. Montine, E. Coto, E. Boerwinkle, R. C. Petersen, V. Alvarez, F. Rivadeneira, E. M. Reiman, M. Gallo, C. J. O’Donnell, J. S. Reisch, A. C. Bruni, D. R. Royall, M. Dichgans, M. Sano, D. Galimberti, P. St George-Hyslop, E. Scarpini, D. W. Tsuang, M. Mancuso, U. Bonuccelli, A. R. Winslow, A. Daniele, C.-K. Wu, GERAD/PERADES, Charge, ADGC, Eadi, O. Peters, B. Nacmias, M. Riemenschneider, R. Heun, C. Brayne, D. C. Rubinsztein, J. Bras, R. Guerreiro, A. Al-Chalabi, C. E. Shaw, J. Collinge, D. Mann, M. Tsolaki, J. Clarimón, R. Sussams, S. Lovestone, M. C. O’Donovan, M. J. Owen, T. W. Behrens, S. Mead, A. M. Goate, A. G. Uitterlinden, C. Holmes, C. Cruchaga, M. Ingelsson, D. A. Bennett, J. Powell, T. E. Golde, C. Graff, P. L. De Jager, K. Morgan, N. Ertekin-Taner, O. Combarros, B. M. Psaty, P. Passmore, S. G. Younkin, C. Berr, V. Gudnason, D. Rujescu, D. W. Dickson, J.-F. Dartigues, A. L. DeStefano, S. Ortega-Cubero, H. Hakonarson, D. Campion, M. Boada, J. K. Kauwe, L. A. Farrer, C. Van Broeckhoven, M. A. Ikram, L. Jones, J. L. Haines, C. Tzourio, L. J. Launer, V. Escott-Price, R. Mayeux, J.-F. Deleuze, N. Amin, P. A. Holmans, M. A. Pericak-Vance, P. Amouyel, C. M. van Duijn, A. Ramirez, L.-S. Wang, J.-C. Lambert, S. Seshadri, J. Williams, G. D. Schellenberg, Rare coding variants in PLCG2, ABI3, and TREM2 implicate microglial-mediated innate immunity in Alzheimer’s disease. Nat. Genet. 49, 1373–1384 (2017).

24. C. Bellenguez, F. Küçükali, I. E. Jansen, L. Kleineidam, S. Moreno-Grau, N. Amin, A. C. Naj, R. Campos-Martin, B. Grenier-Boley, V. Andrade, P. A. Holmans, A. Boland, V. Damotte, S. J. van der Lee, M. R. Costa, T. Kuulasmaa, Q. Yang, I. de Rojas, J. C. Bis, A. Yaqub, I. Prokic, J. Chapuis, S. Ahmad, V. Giedraitis, D. Aarsland, P. Garcia-Gonzalez, C. Abdelnour, E. Alarcón-Martín, D. Alcolea, M. Alegret, I. Alvarez, V. Álvarez, N. J. Armstrong, A. Tsolaki, C. Antúnez, I. Appollonio, M. Arcaro, S. Archetti, A. A. Pastor, B. Arosio, L. Athanasiu, H. Bailly, N. Banaj, M. Baquero, S. Barral, A. Beiser, A. B. Pastor, J. E. Below, P. Benchek, L. Benussi, C. Berr, C. Besse, V. Bessi, G. Binetti, A. Bizarro, R. Blesa, M. Boada, E. Boerwinkle, B. Borroni, S. Boschi, P. Bossù, G. Bråthen, J. Bressler, C. Bresner, H. Brodaty, K. J. Brookes, L. I. Brusco, D. Buiza-Rueda, K. Bûrger, V. Burholt, W. S. Bush, M. Calero, L. B. Cantwell, G. Chene, J. Chung, M. L. Cuccaro, Á. Carracedo, R. Cecchetti, L. Cervera-Carles, C. Charbonnier, H.-H. Chen, C. Chillotti, S. Ciccone, J. A. H. R. Claassen, C. Clark, E. Conti, A. Corma-Gómez, E. Costantini, C. Custodero, D. Daian, M. C. Dalmasso, A. Daniele, E. Dardiotis, J.-F. Dartigues, P. P. de Deyn, K. de Paiva Lopes, L. D. de Witte, S. Debette, J. Deckert, T. del Ser, N. Denning, A. DeStefano, M. Dichgans, J. Diehl-Schmid, M. Diez-Fairen, P. D. Rossi, S. Djurovic, E. Duron, E. Düzel, C. Dufouil, G. Eiriksdottir, S. Engelborghs, V. Escott-Price, A. Espinosa, M. Ewers, K. M. Faber, T. Fabrizio, S. F. Nielsen, D. W. Fardo, L. Farotti, C. Fenoglio, M. Fernández-Fuertes, R. Ferrari, C. B. Ferreira, E. Ferri, B. Fin, P. Fischer, T. Fladby, K. Fließbach, B. Fongang, M. Fornage, J. Fortea, T. M. Foroud, S. Fostinelli, N. C. Fox, E. Franco-Macías, M. J. Bullido, A. Frank-García, L. Froelich, B. Fulton-Howard, D. Galimberti, J. M. García-Alberca, P. García-González, S. Garcia-Madrona, G. Garcia-Ribas, R. Ghidoni, I. Giegling, G. Giorgio, A. M. Goate, O. Goldhardt, D. Gomez-Fonseca, A. González-Pérez, C. Graff, G. Grande, E. Green, T. Grimmer, E. Grünblatt, M. Grunin, V. Gudnason, T. Guetta-Baranes, A. Haapasalo, G. Hadjigeorgiou, J. L. Haines, K. L. Hamilton-Nelson, H. Hampel, O. Hanon, J. Hardy, A. M. Hartmann, L. Hausner, J. Harwood, S. Heilmann-Heimbach, S. Helisalmi, M. T. Heneka, I. Hernández, M. J. Herrmann, P. Hoffmann, C. Holmes, H. Holstege, R. H. Vilas, M. Hulsman, J. Humphrey, G. J. Biessels, X. Jian, C. Johansson, G. R. Jun, Y. Kastumata, J. Kauwe, P. G. Kehoe, L. Kilander, A. K. Ståhlbom, M. Kivipelto, A. Koivisto, J. Kornhuber, M. H. Kosmidis, W. A. Kukull, P. P. Kuksa, B. W. Kunkle, A. B. Kuzma, C. Lage, E. J. Laukka, L. Launer, A. Lauria, C.-Y. Lee, J. Lehtisalo, O. Lerch, A. Lleó, W. Longstreth, O. Lopez, A. L. de Munain, S. Love, M. Löwemark, L. Luckcuck, K. L. Lunetta, Y. Ma, J. Macías, C. A. MacLeod, W. Maier, F. Mangialasche, M. Spallazzi, M. Marquié, R. Marshall, E. R. Martin, A. M. Montes, C. M. Rodríguez, C. Masullo, R. Mayeux, S. Mead, P. Mecocci, M. Medina, A. Meggy, S. Mehrabian, S. Mendoza, M. Menéndez-González, P. Mir, S. Moebus, M. Mol, L. Molina-Porcel, L. Montrreal, L. Morelli, F. Moreno, K. Morgan, T. Mosley, M. M. Nöthen, C. Muchnik, S. Mukherjee, B. Nacmias, T. Ngandu, G. Nicolas, B. G. Nordestgaard, R. Olaso, A. Orellana, M. Orsini, G. Ortega, A. Padovani, C. Paolo, G. Papenberg, L. Parnetti, F. Pasquier, P. Pastor, G. Peloso, A. Pérez-Cordón, J. Pérez-Tur, P. Pericard, O. Peters, Y. A. L. Pijnenburg, J. A. Pineda, G. Piñol-Ripoll, C. Pisanu, T. Polak, J. Popp, J. Priller, R. Puerta, O. Quenez, I. Quintela, J. Q. Thomassen, A. Rábano, I. Rainero, F. Rajabli, I. Ramakers, L. M. Real, M. J. T. Reinders, C. Reitz, D. Reyes-Dumeyer, P. Ridge, S. Riedel-Heller, P. Riederer, N. Roberto, E. Rodriguez-Rodriguez, A. Rongve, I. R. Allende, M. Rosende-Roca, J. L. Royo, E. Rubino, D. Rujescu, M. E. Sáez, P. Sakka, I. Saltvedt, Á. Sanabria, M. B. Sánchez-Arjona, F. Sanchez-Garcia, P. S. Juan, R. Sánchez-Valle, S. B. Sando, C. Sarnowski, C. L. Satizabal, M. Scamosci, N. Scarmeas, E. Scarpini, P. Scheltens, N. Scherbaum, M. Scherer, M. Schmid, A. Schneider, J. M. Schott, G. Selbæk, D. Seripa, M. Serrano, J. Sha, A. A. Shadrin, O. Skrobot, S. Slifer, G. J. L. Snijders, H. Soininen, V. Solfrizzi, A. Solomon, Y. Song, S. Sorbi, O. Sotolongo-Grau, G. Spalletta, A. Spottke, A. Squassina, E. Stordal, J. P. Tartan, L. Tárraga, N. Tesí, A. Thalamuthu, T. Thomas, G. Tosto, L. Traykov, L. Tremolizzo, A. Tybjærg-Hansen, A. Uitterlinden, A. Ullgren, I. Ulstein, S. Valero, O. Valladares, C. Van Broeckhoven, J. Vance, B. N. Vardarajan, A. van der Lugt, J. Van Dongen, J. van Rooij, J. van Swieten, R. Vandenberghe, F. Verhey, J.-S. Vidal, J. Vogelgsang, M. Vyhnalek, M. Wagner, D. Wallon, L.-S. Wang, R. Wang, L. Weinhold, J. Wiltfang, G. Windle, B. Woods, M. Yannakoulia, H. Zare, Y. Zhao, X. Zhang, C. Zhu, M. Zulaica, L. A. Farrer, B. M. Psaty, M. Ghanbari, T. Raj, P. Sachdev, K. Mather, F. Jessen, M. A. Ikram, A. de Mendonça, J. Hort, M. Tsolaki, M. A. Pericak-Vance, P. Amouyel, J. Williams, R. Frikke-Schmidt, J. Clarimon, J.-F. Deleuze, G. Rossi, S. Seshadri, O. A. Andreassen, M. Ingelsson, M. Hiltunen, K. Sleegers, G. D. Schellenberg, C. M. van Duijn, R. Sims, W. M. van der Flier, A. Ruiz, A. Ramirez, J.-C. Lambert, New insights into the genetic etiology of Alzheimer’s disease and related dementias. Nat. Genet. 54, 412–436 (2022).

25. D. V. Hansen, J. E. Hanson, M. Sheng, Microglia in Alzheimer’s disease. J. Cell Biol. 217, 459–472 (2018).

26. C. Condello, P. Yuan, A. Schain, J. Grutzendler, Microglia constitute a barrier that prevents neurotoxic protofibrillar Aβ42 hotspots around plaques. Nat. Commun. 6, 6176 (2015).

27. P. Yuan, C. Condello, C. D. Keene, Y. Wang, T. D. Bird, S. M. Paul, W. Luo, M. Colonna, D. Baddeley, J. Grutzendler, TREM2 Haplodeficiency in Mice and Humans Impairs the Microglia Barrier Function Leading to Decreased Amyloid Compaction and Severe Axonal Dystrophy. Neuron 92, 252–264 (2016).

28. F. L. Yeh, Y. Wang, I. Tom, L. C. Gonzalez, M. Sheng, TREM2 Binds to Apolipoproteins, Including APOE and CLU/APOJ, and Thereby Facilitates Uptake of Amyloid-Beta by Microglia. Neuron 91, 328–340 (2016).

29. W. M. Song, S. Joshita, Y. Zhou, T. K. Ulland, S. Gilfillan, M. Colonna, Humanized TREM2 mice reveal microglia-intrinsic and -extrinsic effects of R47H polymorphism. J. Exp. Med. 215, 745–760 (2018).

30. H. Keren-Shaul, A. Spinrad, A. Weiner, O. Matcovitch-Natan, R. Dvir-Szternfeld, T. K. Ulland, E. David, K. Baruch, D. Lara-Astaiso, B. Toth, S. Itzkovitz, M. Colonna, M. Schwartz, I. Amit, A Unique Microglia Type Associated with Restricting Development of Alzheimer’s Disease. Cell 169, 1276–1290.e17 (2017).

31. K. Srinivasan, B. A. Friedman, A. Etxeberria, M. A. Huntley, M. P. van der Brug, O. Foreman, J. S. Paw, Z. Modrusan, T. G. Beach, G. E. Serrano, D. V. Hansen, Alzheimer’s Patient Microglia Exhibit Enhanced Aging and Unique Transcriptional Activation. Cell Rep. 31, 107843 (2020).

32. K. Cosker, A. Mallach, J. Limaye, T. M. Piers, J. Staddon, S. J. Neame, J. Hardy, J. M. Pocock, Microglial signalling pathway deficits associated with the patient derived R47H. TREM2 variants linked to AD indicate inability to activate inflammasome. Sci. Rep. 11, 13316 (2021).

33. Y. Zhou, W. M. Song, P. S. Andhey, A. Swain, T. Levy, K. R. Miller, P. L. Poliani, M. Cominelli, S. Grover, S. Gilfillan, M. Cella, T. K. Ulland, K. Zaitsev, A. Miyashita, T. Ikeuchi, M. Sainouchi, A. Kakita, D. A. Bennett, J. A. Schneider, M. R. Nichols, S. A. Beausoleil, J. D. Ulrich, D. M. Holtzman, M. N. Artyomov, M. Colonna, Human and mouse single-nucleus transcriptomics reveal TREM2-dependent and TREM2-independent cellular responses in Alzheimer’s disease. Nat. Med. 26, 131–142 (2020).

34. A. P. Tsai, C. Dong, P. B.-C. Lin, A. L. Oblak, G. Viana Di Prisco, N. Wang, N. Hajicek, A. J. Carr, E. K. Lendy, O. Hahn, M. Atkins, A. G. Foltz, J. Patel, G. Xu, M. Moutinho, J. Sondek, Q. Zhang, A. D. Mesecar, Y. Liu, B. K. Atwood, T. Wyss-Coray, K. Nho, S. J. Bissel, B. T. Lamb, G. E. Landreth, Genetic variants of phospholipase C-γ2 alter the phenotype and function of microglia and confer differential risk for Alzheimer’s disease. Immunity 56, 2121–2136.e6 (2023).

35. Z. Yin, N. Rosenzweig, K. L. Kleemann, X. Zhang, W. Brandão, M. A. Margeta, C. Schroeder, K. N. Sivanathan, S. Silveira, C. Gauthier, D. Mallah, K. M. Pitts, A. Durao, S. Herron, H. Shorey, Y. Cheng, J.-L. Barry, R. K. Krishnan, S. Wakelin, J. Rhee, A. Yung, M. Aronchik, C. Wang, N. Jain, X. Bao, E. Gerrits, N. Brouwer, A. Deik, D. G. Tenen, T. Ikezu, N. G. Santander, G. L. McKinsey, C. Baufeld, D. Sheppard, S. Krasemann, R. Nowarski, B. J. L. Eggen, C. Clish, R. E. Tanzi, C. Madore, T. D. Arnold, D. M. Holtzman, O. Butovsky, APOE4 impairs the microglial response in Alzheimer’s disease by inducing TGFβ-mediated checkpoints. Nat. Immunol. 24, 1839–1853 (2023).

36. R. Bruderer, O. M. Bernhardt, T. Gandhi, Y. Xuan, J. Sondermann, M. Schmidt, D. Gomez-Varela, L. Reiter, Optimization of Experimental Parameters in Data-Independent Mass Spectrometry Significantly Increases Depth and Reproducibility of Results. Mol. Cell. Proteomics 16, 2296–2309 (2017).

37. M. Tognetti, K. Sklodowski, S. Müller, D. Kamber, J. Muntel, R. Bruderer, L. Reiter, Biomarker Candidates for Tumors Identified from Deep-Profiled Plasma Stem Predominantly from the Low Abundant Area. J. Proteome Res. 21, 1718–1735 (2022).

38. T. D. Wu, S. Nacu, Fast and SNP-tolerant detection of complex variants and splicing in short reads. Bioinformatics 26, 873–881 (2010).

39. S. J. Fleming, M. D. Chaffin, A. Arduini, A.-D. Akkad, E. Banks, J. C. Marioni, A. A. Philippakis, P. T. Ellinor, M. Babadi, Unsupervised removal of systematic background noise from droplet-based single-cell experiments using CellBender. Nat. Methods 20, 1323–1335 (2023).

40. M. D. Robinson, D. J. McCarthy, G. K. Smyth, edgeR: a Bioconductor package for differential expression analysis of digital gene expression data. Bioinformatics 26, 139–140 (2010).

41. C. W. Law, Y. Chen, W. Shi, G. K. Smyth, voom: Precision weights unlock linear model analysis tools for RNA-seq read counts. Genome Biol. 15, R29 (2014).

42. B. A. Friedman, K. Srinivasan, G. Ayalon, W. J. Meilandt, H. Lin, M. A. Huntley, Y. Cao, S.-H. Lee, P. C. G. Haddick, H. Ngu, Z. Modrusan, J. L. Larson, J. S. Kaminker, M. P. van der Brug, D. V. Hansen, Diverse Brain Myeloid Expression Profiles Reveal Distinct Microglial Activation States and Aspects of Alzheimer’s Disease Not Evident in Mouse Models. Cell Rep. 22, 832–847 (2018).

43. Scientific Image and Illustration Software. https://www.biorender.com/.

